# Recovery Trajectories in Post-stroke Ataxia: Distinct Longitudinal Classes Characterized by Initial Severity and Recovery Timing

**DOI:** 10.64898/2026.03.10.26348027

**Authors:** Yuichiro Yamasaki, Yusaku Takamura, Hirofumi Sato, Katsunobu Okuma, Yohei Kobayashi, Ariyasu Kamijima, Shinjiro Takaishi, Hideyuki Maruki, Shu Morioka

## Abstract

**Purpose:** Longitudinal recovery from post-stroke ataxia remains poorly characterized. We examined whether recovery could be characterized by model-derived initial severity (*α*), asymptotic proportional recovery coefficient (*r*), and recovery time constant (*τ*), and whether trajectories differed across these parameters.

**Methods:** Longitudinal Scale for the Assessment and Rating of Ataxia (SARA) scores from 80 patients with post-stroke ataxia were analyzed using a Bayesian nonlinear mixed-effects model. Trajectory classes were identified using a latent class mixed model. Walking independence was analyzed with interval censoring. Posterior-propagated regressions included *α*, age, sex, and Trail Making Test Part A (TMT-A), with baseline Berg Balance Scale (BBS) included in a functional sensitivity model.

**Results:** Three trajectory classes were identified (n=55, 19, and 6). Class 1 had lower initial severity and shorter τ. Class 2 had intermediate initial severity and τ broadly comparable to that of Class 1. Class 3 had the greatest initial severity and markedly longer τ. Posterior summaries of r overlapped, whereas model-implied proportional improvement by 180 days differed across classes (approximately 0.80, 0.78, and 0.53). Walking independence was observed in 52/55, 18/19, and 0/6 participants. A 10-point higher baseline BBS score was associated with a 15.2-day shorter τ (95% credible interval, −27.3 to −5.2). Poorer TMT-A performance was directionally associated with longer τ, with substantial uncertainty; associations with r remained uncertain.

**Conclusions:** Recovery trajectories were heterogeneous, particularly in initial severity and timing. Separating recovery magnitude from timing may be more clinically informative than initial severity alone. External validation is needed before patient-level prognostic use.

## Introduction

Post-stroke ataxia can substantially restrict balance, walking, and activities of daily living through impaired coordination and postural control.^1–4^ Cerebellar infarction accounts for approximately 3.4% of ischemic strokes, and cerebellar hemorrhage for approximately 10% of intracerebral hemorrhages.^5,6^ Ataxia may also arise from brainstem lesions involving cerebellar pathways.^2,7,8^ Although functional improvement and domain-specific changes have been reported over weeks to months,^9–12^ the temporal course of recovery from post-stroke ataxia has not been adequately quantified. The proportional recovery rule describes improvement as a proportion of initial motor impairment and has been examined primarily in limb paresis after stroke.^13–15^ In the original upper-limb study of ischemic stroke, this relationship did not characterize all patients with severe initial impairment.^13^ Subsequent studies linked recovery patterns to corticospinal integrity.^16,17^ Longitudinal modeling has characterized nonlinear, time-dependent post-stroke motor recovery trajectories.^18,19^ Exponential trajectories describe relatively rapid early improvement followed by a gradual approach to an asymptote, using three conceptually distinct parameters: model-estimated initial severity (α), asymptotic proportional recovery coefficient (r), and recovery time constant (τ).^18,19^ The clinical relevance of this decomposition for post-stroke ataxia cannot be inferred directly from studies of paresis. Recovery from motor paresis is closely associated with the integrity of descending corticospinal pathways,^16,17^ whereas ataxia may result from lesions of the cerebellum or cerebellar pathways within the brainstem^2,7,8^. Cerebellar lesions may also produce cognitive-affective dysfunction, including executive and visuospatial deficits.^20^ These features provide a rationale for considering broader cognitive and motor-task performance when examining recovery dynamics.^21^ We therefore examined two hypotheses: (i) recovery from post-stroke ataxia cannot be adequately characterized by initial severity alone and could be more fully characterized by model-estimated initial severity, asymptotic proportional improvement, and recovery time scale; and (ii) recovery magnitude and timing may vary across patients as distinct dimensions, with interindividual variation expressed as multiple latent longitudinal trajectories of Scale for the Assessment and Rating of Ataxia (SARA) scores with different α, r, and τ profiles rather than a single population-average pattern. We estimated these parameters using a Bayesian nonlinear mixed-effects model and identified classes directly from repeated SARA scores using a latent class mixed model. We further explored observed walking independence across classes and clinical characteristics associated with the model-derived parameters.

In this study, recovery refers to longitudinal change in ataxia-related impairment as captured by the SARA.^22,23^ Neurological restitution denotes restoration of impaired function, whereas behavioral compensation denotes the use of alternative strategies to achieve task goals.^24^ Because changes in the SARA total score cannot directly distinguish the contributions of neurological restitution and behavioral compensation,^24^ *α*, *r*, and *τ* were interpreted as parameters describing clinically and temporally distinct features of the observed SARA trajectory rather than specific recovery mechanisms.

## Methods

### Participants

From June 2020 to April 2025, we screened 174 patients with stroke with ataxia admitted to convalescent rehabilitation wards at three collaborative research facilities. Inclusion criteria were as follows: (1) stroke involving the cerebellum and/or brainstem within the vertebrobasilar territory; (2) presence of ataxia confirmed by a neurologist using the finger-to-finger, finger-to-nose, and heel-to-knee tests; and (3) Brunnstrom recovery stage V or higher.^3^ Neurologists at each facility confirmed stroke localization using magnetic resonance imaging or computed tomography performed either during the acute phase or after transfer to the rehabilitation hospital. Of the 174 screened patients, 155 met the initial eligibility criteria and were enrolled. Primary reasons for exclusion at this stage included interruption of rehabilitation (n = 4), orthopedic lower limb pain (n = 5), severe sensory disturbance (e.g., loss of thermal or pain sensation; n = 5), and inability to undergo evaluation at admission (n = 5). Subsequently, patients were excluded from the analysis because of either cognitive impairment (Japanese version of the Mini-Mental State Examination [MMSE-J] score ≤23; n = 30), which could compromise the reliability of repeated SARA assessments, or insufficient longitudinal data (fewer than three assessments; n = 45). Ultimately, 80 patients (36 male, 44 female; mean age 69.2 ± 13.2 years) were included in the final analysis. The participant selection process is shown in Supplementary Figure S1, and baseline characteristics are presented in Table 1. This study was conducted with the approval of the ethics committees of the Maruki Memorial Medical and Social Welfare Center, Saitama Citizens Medical Center, and Saitama Sekishinkai Hospital (approval numbers: Maruki Memorial Medical and Social Welfare Center: 32; Saitama Citizens Medical Center: 2020-09; Saitama Sekishinkai Hospital: 2023-5). All participants provided written informed consent in accordance with the Declaration of Helsinki.

**Table 1.** Participant characteristics and clinical measures according to LCMM-derived trajectory class. Data are presented as absolute numbers for categorical variables and median [interquartile range] for continuous variables. Clinical scale scores are admission measurements, with one value per participant. Scores were available for all 80 participants (Class 1, n=55; Class 2, n=19; Class 3, n=6). Physical therapy time was calculated for each participant from the recorded mean number of PT units per treatment day, using 20 minutes per unit. The table presents the median [interquartile range] of these participant-level mean values. BMI, body mass index; mRS, modified Rankin Scale; SARA, Scale for the Assessment and Rating of Ataxia; BBS, Berg Balance Scale; Mini-BESTest, Mini Balance Evaluation Systems Test; ABMS-Ⅱ, Ability for Basic Movement Scale Ⅱ; m-FIM, motor Functional Independence Measure; c-FIM, cognitive Functional Independence Measure; MMSE-J, Japanese version of the Mini-Mental State Examination; TMT-A, Trail Making Test Part A; FAB, Frontal Assessment Battery; LCMM, latent class mixed model.

| Basic attributes and clinical indicators | All patients (n=80) | Class 1 (n=55) | Class 2 (n=19) | Class 3 (n=6) |
| --- | --- | --- | --- | --- |
| Age (years) | 72.5 [58.8, 79.0] | 73.0 [58.5, 79.0] | 72.0 [58.0, 77.5] | 74.0 [64.0, 81.0] |
| Sex: male/ female | 36 / 44 | 26 / 29 | 7 / 12 | 3 / 3 |
| BMI (kg/m<sup>2</sup>) | 22.1 [20.0, 24.5] | 22.5 [20.2, 24.7] | 22.0 [19.9, 24.3] | 21.5 [19.5, 21.7] |
| Stroke type: hemorrhages/ infarcts | 25 / 55 | 13/ 42 | 8/ 11 | 4/ 2 |
| Affected side: right/ left/ bilateral | 49 / 30 / 1 | 31/ 23/ 1 | 15/ 4/ 0 | 3/ 3/ 0 |
| Premorbid mRS: 0/ 1/ 2/ 3 | 72 / 6 / 1 / 1 | 50/ 3/ 1/ 1 | 16/ 3/ 0/ 0 | 6/ 0/ 0/ 0 |
| Lesion site<br>(cerebellum/midbrain/pons/medulla oblongata/multiple lesions) | 49 / 1 / 14 / 11 / 5 | 27/ 1/ 12/ 10/ 5 | 16/ 0/ 2/ 1/ 0 | 6/ 0/ 0/ 0/ 0 |
| Time since stroke (days) | 20.5 [13.0, 29.5] | 18 [12.0, 24.0] | 38 [19.0, 42.5] | 25 [21.0, 35.8] |
| Length of stay (days) | 70.0 [61.8, 91.0] | 67 [57.5, 82.5] | 73 [66.5, 87.0] | 122.5 [119.0, 123.0] |
| Mean physical therapy time (min per treatment day) | 62.3 [60.0, 72.7] | 67.6 [60.6, 77.1] | 61.2 [58.0, 62.8] | 61.0 [59.5, 62.7] |
| SARA | 11.5 [8.50, 14.1] | 9.50 [6.50, 12.0] | 14.5 [13.8, 19.0] | 27.5 [27.0, 28.0] |
| BBS | 31.0 [24.0, 40.0] | 35.0 [28.0, 42.5] | 24.0 [18.0, 27.5] | 7.00 [7.00, 9.25] |
| Mini-BESTest | 7.50 [2.00, 14.3] | 11.0 [5.50, 17.0] | 2.00 [1.00, 4.50] | 1.00 [1.00, 1.00] |
| ABMS-II | 26.0 [21.0, 28.0] | 27.0 [23.5, 28.0] | 21.0 [21.0, 23.0] | 16.0 [16.0, 16.8] |
| m-FIM | 51.5 [39.8, 62.0] | 54.0 [48.5, 65.5] | 40.0 [34.5, 44.0] | 20.0 [17.5, 21.0] |
| c-FIM | 30.0 [26.0, 33.0] | 30.0 [26.5, 33.0] | 30.0 [27.5, 30.0] | 25.0 [25.0, 25.8] |
| MMSE-J | 27.0 [25.0, 30.0] | 28.0 [26.0, 30.0] | 26.0 [24.0, 27.0] | 24.5 [24.0, 25.0] |
| TMT-A (s) | 71.7 [49.0, 109.7] | 69.0 [43.8, 101.2] | 68.0 [57.9, 105.6] | 190.0 [109.8, 216.5] |
| FAB | 15.0 [10.8, 16.0] | 15.0 [12.0, 17.0] | 12.0 [10.0, 15.5] | 10.0 [9.25, 11.5] |

### Clinical Measurements

The following demographic and clinical data were extracted from medical records: age at stroke onset, sex, pre-stroke modified Rankin Scale score, stroke type, lesion site, ataxia side, time from onset to admission, and length of hospital stay, as well as mean physical therapy time per treatment day during hospitalization. Assessments of motor, cognitive, and higher brain functions were performed at admission and subsequently reassessed at 30-day intervals. The pre-stroke modified Rankin Scale assesses baseline functional status on a 6-point scale from grades 0 to 5.^25^ The SARA quantifies ataxia severity, with scores ranging from 0 (no symptoms) to 40 (most severe).^22^ The Berg Balance Scale (BBS) assesses balance ability on a scale of 0–56, with higher scores indicating better function.^26^ The Mini-Balance Evaluation Systems Test also evaluates dynamic balance (maximum score 28; higher scores indicate better balance).^27^ The Ability for Basic Movement Scale-II quantifies the assistance required for basic mobility on a scale of 5–30.^28^ The Functional Independence Measure (FIM) comprises 18 items (13 motor and 5 cognitive) to assess independence in daily activities, with a total score of 18–126; higher scores indicate greater independence.^29^ The FIM locomotion item was recorded in walking mode on a 1–7 scale, with scores ≥6 indicating walking independence. It was assessed at admission, at approximately 30-day intervals, and at discharge. The Japanese version of the Mini-Mental State Examination is a screening test for cognitive function (maximum score 30).^30^ Trail Making Test Part A (TMT-A) completion time was used as a timed measure reflecting visual search, motor speed, and attentional performance and was performed using the dominant hand (or the nondominant hand if the former was affected), with longer completion times indicating poorer performance.^31^ Because completion time may also reflect visuomotor performance, TMT-A was not interpreted as a pure measure of attention. No separate accuracy/error score was collected, and no participant discontinued TMT-A because of fatigue. The Frontal Assessment Battery (FAB) evaluates frontal lobe function (total score 18), and its clinical utility has been established in patients with stroke.^32^ For the present analyses, FAB was used as an alternative cognitive measure in the sensitivity analysis of cognition. Comprehensive physical therapy (PT)^33^ was provided from admission to discharge, including range of motion exercises, muscle strengthening, trunk and limb coordination exercises, balance training, and gait training based on protocols described by Miyai^34^ and Ilg.^35^ For each participant, mean PT time per treatment day was calculated by multiplying the recorded mean PT units per treatment day by 20 minutes. The denominator was PT treatment days, not total hospital days.

### Statistical Analyses

#### Estimation of Recovery Trajectories Using Bayesian Nonlinear Mixed-effects Models

Longitudinal SARA scores were modeled using a Bayesian nonlinear mixed-effects exponential decay model, which assumed rapid early improvement that gradually slowed. For patient i at time j:

##### 1. Observation Model

SARA scores were assumed normally distributed.

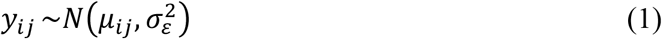

##### 2. Structural Model

The expected value (*μij*) is defined by the exponential decay function.

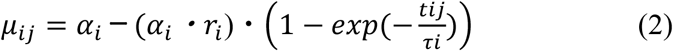

Here, tij represents the time since onset (days). The individual model parameters are defined as follows:

*αᵢ*: Model-estimated initial SARA severity (0 = no ataxia; 40 = maximum SARA severity); higher values indicate greater initial ataxia severity.

*rᵢ*: Proportional recovery coefficient (0 = no model-implied recovery; 1 = complete asymptotic recovery toward SARA score of 0); higher values indicate a greater proportion of the initial SARA severity recovered asymptotically.

*τᵢ*: Recovery time constant (>0 days); at t = τᵢ, approximately 63.2% of the model-implied recoverable change has occurred, with larger values indicating slower recovery. The α-r-τ parameterization was adapted from prior longitudinal stroke-recovery modeling.^19^

##### 3. Parameter Transformation and Link Functions

To enforce parameter bounds, *αᵢ*, *rᵢ*, and *τᵢ* were linked to participant-specific internal predictors *aᵢ*, *pᵢ*, and *λᵢ* as follows:

A. Initial severity (*αᵢ*): range 0–40.

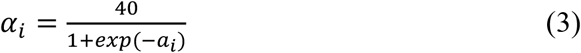
B. Proportional recovery coefficient (*rᵢ*): range 0–1.

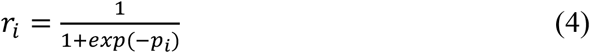
C. Recovery time constant (*τᵢ*): >0 days; A constant of 0.142 days (approximately 1/7 day) was added for computational stability.

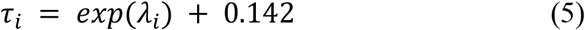

##### 4. Hierarchical Structure and Estimation

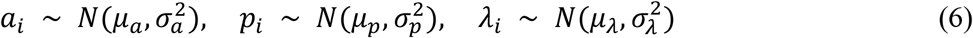

Separate population-level intercepts and between-participant random-effect variances were estimated for the internal predictors *aᵢ*, *pᵢ*, and *λᵢ*; cross-parameter random-effect covariances were not estimated. Parameter estimation was performed using Markov chain Monte Carlo (MCMC) with weakly informative priors. The population-level intercepts for aᵢ and pᵢ were assigned Normal (0, 2) priors, whereas that for *λᵢ* was assigned a Normal (3, 1) prior; the second argument in these Normal prior specifications denotes the standard deviation. The corresponding random-effect standard deviations (SDs) were assigned half-Cauchy priors with scale parameters of 2, 1, and 1, respectively, and the residual SD was assigned a half-Cauchy prior with a scale parameter of 5. These priors were not calibrated to parameter estimates from previous studies but were selected as weakly informative distributions on their respective model scales. Four MCMC chains of 4,000 iterations (2,000 warm-up) were run with adapt_delta = 0.99 and max_treedepth = 15. Sampling was assessed using R-hat (≤1.01), bulk and tail effective sample sizes, Monte Carlo standard errors (MCSEs) of posterior means, and trace plots; model fit was assessed using posterior predictive checks³⁶ (Supplementary Figure S5; Supplementary Table S7).

##### 5. Model-Implied Proportional Improvement by 180 Days

Model-implied proportional improvement by 180 days after stroke onset was defined as follows:

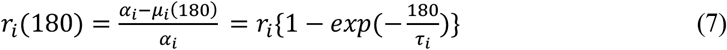

Here, *μᵢ*(180) denotes the model-implied expected SARA score at day 180. The derived quantity *rᵢ*(180) represents the proportion of estimated initial SARA severity recovered by day 180 under the fitted model, rather than an observed day-180 change score.

##### 6. Model Validation and Sensitivity Analyses

Practical identifiability was assessed by posterior dependence, parameter-recovery simulations reproducing the observed assessment schedule, and wider-prior sensitivity³⁷,³⁸ (Supplementary Figures S2–S3; Supplementary Tables S1–S2). Supplementary Methods, Sections 1–2, provide details of data generation, simulation-refit acceptance criteria, and the original and wider priors. Model-form sensitivity compared the single-exponential model with linear, bi-exponential, logistic, Gompertz, and spline alternatives using the same participant-grouped five-fold partition. In each fold, models were fitted to data from 64 participants (80%) and evaluated on the complete longitudinal series of 16 held-out participants (20%); each participant was held out once across the five folds.

Held-out participants were treated as new patients, with random effects sampled from the distribution implied by each training fit. The primary measure was participant-level joint expected log predictive density (ELPD) on the original SARA scale. Root mean squared error (RMSE), predictive interval coverage, and full-data trajectory and residual assessments were complementary. Computational details and data-dependent settings retained across folds are provided in Supplementary Methods, Section 3 (Supplementary Table S3; Supplementary Figure S4).

#### Latent Class Trajectory Analysis

Trajectory classes were estimated directly from longitudinal SARA observations using latent class mixed models (LCMMs) rather than Bayesian parameter point estimates.^39^ This approach has recently been applied to a large multicenter spinal cord injury cohort.^40^ Linear random-intercept and quadratic common-curvature Gaussian LCMMs with an identity link were initially compared with one to three classes and subsequently extended to four classes as a sensitivity analysis. Bayesian information criterion (BIC) was the primary selection criterion, supplemented by ICL, SABIC, classification quality, class size, and optimization reproducibility; AIC was additionally reported in Supplementary Table S4.

SARA scores were standardized, and time since stroke onset was centered on the median first-assessment day across participants and expressed in 30-day units. Participants were assigned to the class with the highest posterior membership probability. Class reproducibility was assessed using participant-level bootstrap resampling, with optimization status evaluated separately from the availability of a usable fitted solution (Figure 1; Supplementary Tables S4–S5). Bayesian estimates of *α*, *r*, and *τ* and the derived quantity *r*(180), defined in Equation (7), were used only to characterize classes after assignment. Class membership was held fixed for these summaries. Computational details are provided in Supplementary Methods, under “LCMM estimation and class-specific summaries.”

**Figure 1.**
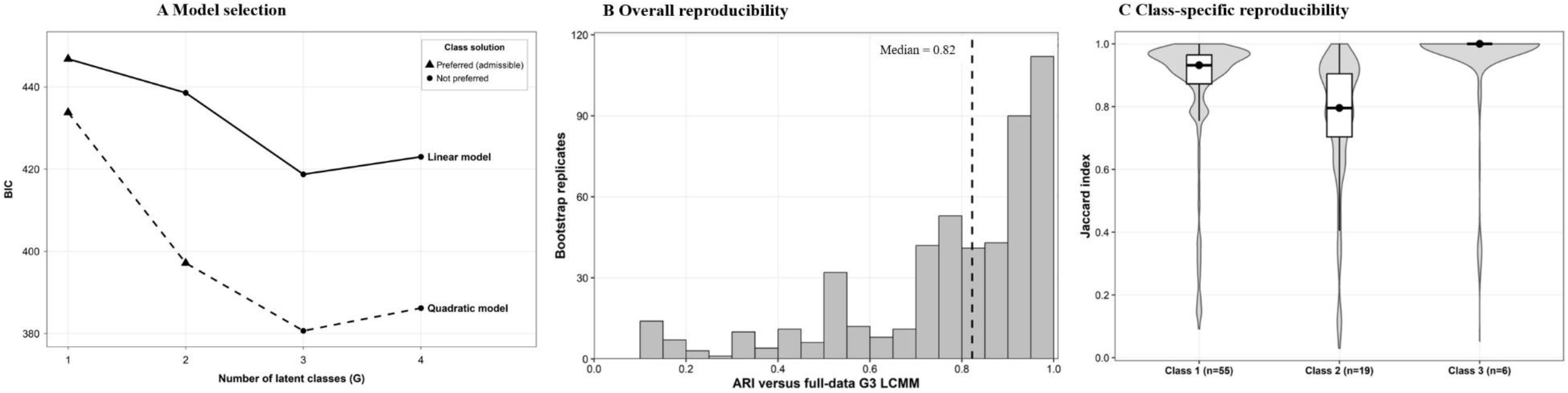
Selection and reproducibility of LCMM-derived recovery trajectory classes. (A) Bayesian information criterion (BIC) across one- to four-class linear random-intercept and quadratic common-curvature random-intercept LCMMs. Four-class candidates were added as a sensitivity analysis. Lower BIC indicates better penalized model fit. (B) Distribution of adjusted Rand indices (ARIs) comparing classifications of the original participants under the bootstrap-fitted models with the full-data reference classification. All successful bootstrap fits were included, irrespective of optimization-resolution status. The dashed line indicates the median ARI. (C) Class-specific Jaccard index distributions from the same bootstrap replicates. Boxplots indicate the median and interquartile range. Bootstrap procedures and optimization criteria are described in Supplementary Methods. Panels B and C refer only to the retained three-class quadratic common-curvature model; the number and structure of classes were held fixed during bootstrap resampling. Symbols indicate the preferred internal size and classification criteria, not the final model-selection decision. Triangles denote solutions meeting those criteria, and circles denote solutions not meeting all of them; single-class solutions are shown as triangles by convention. BIC, Bayesian information criterion; ARI, adjusted Rand index; LCMM, latent class mixed model.

#### Observed Walking Independence

Walking independence was examined descriptively across trajectory classes derived from the full longitudinal SARA series. The event was defined as the first attainment of a FIM locomotion score ≥6, with stroke onset used as the time origin. Because assessments were conducted intermittently, event times were classified as left-, interval-, or right-censored as appropriate. Class-specific cumulative probabilities of attaining walking independence were estimated using the Turnbull nonparametric maximum likelihood estimator with corresponding 95% confidence intervals.

#### Posterior-Propagated Secondary Analyses

Clinical associations with r and τ were examined using posterior-propagated Gaussian regressions based on 4,000 joint participant-level draws selected from the 8,000 post-warm-up draws of the primary model. The same draw was used for *α* and the outcome parameter, preserving their posterior dependence. For each selected draw and each regression, the residual variance and regression coefficients were sampled from their conditional posterior distribution under the reference prior p(β,σ²) ∝ 1/σ², where σ² denotes the secondary-regression residual variance. Clinical covariates did not feed back into the primary trajectory model. Outcomes were modeled on the logit(*r*) and log(*τ* − 0.142) scales; natural-scale average marginal differences were calculated as detailed in the footnote to Supplementary Table S6. No data-driven variable selection or frequentist P-value testing was used. The core model included natural-scale *α*, age at stroke onset, sex, and TMT-A; the anatomic and functional sensitivity models additionally included stroke subtype and lesion category, and baseline Berg Balance Scale score, respectively. *α* was scaled per 5 SARA points, age per 10 years and treated as time-invariant, and TMT-A was log-transformed and standardized. Admission measurements of TMT-A, FAB, and BBS were used rather than values averaged across assessments. All secondary models included the same 80 participants, for whom data on all required clinical covariates were complete; no imputation was performed.

Supplementary analyses were conducted by replacing TMT-A with FAB, additionally adjusting for mean physical therapy time per PT treatment day during hospitalization, and repeating the core, anatomic, and functional models using the internal initial-severity predictor aᵢ = logit(αᵢ/40) rather than natural-scale α (Supplementary Table S6). As an exploratory magnitude-sensitivity analysis, natural-scale posterior contrasts from the core model and the additional BBS contrast from the functional sensitivity model were compared with regions of practical equivalence (ROPEs) defined as ±0.10 of a reference between-participant standard deviation. The proportions of propagated draws below, within, and above the ROPE were summarized (Supplementary Methods; Supplementary Table S8). Posterior medians and 95% credible intervals remained the primary summaries (Table 3; Figure 3).

#### Sample Size and Software

The study hypotheses and analyses were not preregistered. No a priori sample-size calculation was performed; all eligible participants who met the longitudinal-data requirements were analyzed. Precision and stability were assessed using posterior uncertainty, MCMC diagnostics, parameter-recovery simulations, and patient-level bootstrap resampling rather than post hoc power calculations. Analyses were performed using R version 4.4.2 and the brms/cmdstanr, lcmm, survival, and bayestestR packages.

## Results

### Participant Characteristics and LCMM-Derived Recovery Trajectory Classes

The final cohort comprised 80 participants contributing 283 longitudinal SARA observations. Among the expanded set of eight LCMM candidates, the three-class quadratic common-curvature random-intercept model had the lowest BIC (380.7; four-class counterpart, 386.2; difference, 5.5). ICL also favored three classes, and classification was clearer than in the corresponding four-class model (entropy, 0.879 vs 0.730; mean maximum posterior membership probability, 0.946 vs 0.838). We therefore retained the three-class solution (n=55, 19, and 6) for the primary descriptive analyses (Figure 1; Supplementary Table S4). For the retained three-class model, all 500 patient-level bootstrap replicates completed, with optimization resolved in 491 (98.2%); the median adjusted Rand index was 0.823, and the median Jaccard indices were 0.932, 0.796, and 1.000 for Classes 1-3, respectively (Supplementary Table S5). Median maximum posterior membership probabilities were 0.997, 0.962, and 1.000, respectively. Clinical characteristics are summarized descriptively in Table 1. Admission SARA scores increased across Classes 1–3 (median, 9.5, 14.5, and 27.5), whereas BBS scores decreased (35.0, 24.0, and 7.0).

### Bayesian Nonlinear Model Fit and Validation

Table 2 reports posterior summaries of population-level intercepts and participant-level random-effect SDs. The seven parameters reported in Supplementary Table S7 had R-hat values of 1.000–1.004, with bulk ESSs of 1,083–3,220 and tail ESSs of 2,252–4,930. MCSEs of the posterior means were 1.75–2.94% of the corresponding posterior SDs. Trace plots and the posterior predictive check are shown in Supplementary Figure S5. The corresponding natural-scale population distributions are shown in Supplementary Figure S6.

**Table 2.** Posterior summaries of population-level intercepts and participant-level random-effect standard deviations in the Bayesian nonlinear mixed-effects model. CrI, credible interval; SD, standard deviation; SARA, Scale for the Assessment and Rating of Ataxia. All numerical estimates are reported on the model-internal scales: aᵢ = logit(αᵢ/40), pᵢ = logit(rᵢ), and λᵢ = log(τᵢ − 0.142). Values are posterior means, equal-tailed 95% credible intervals defined by the 2.5th and 97.5th posterior percentiles, and posterior standard deviations. Numerical summaries are rounded to three decimal places. Random-effect SDs quantify between-participant variability on these scales, whereas posterior SDs quantify uncertainty in the estimated parameters. Sampling diagnostics are provided in Supplementary Table S7.

| Section | Parameter | Posterior mean (95% CrI) | Posterior SD |
| --- | --- | --- | --- |
| Population-level intercepts | Initial severity: $\text{logit}(\alpha/40)$ | $-0.275$ ( $-0.546$ to $0.002$ ) | 0.140 |
| Population-level intercepts | Proportional recovery coefficient: $\text{logit}(r)$ | $1.959$ ( $1.535$ to $2.637$ ) | 0.287 |
| Population-level intercepts | Recovery time constant: $\log(\tau - 0.142)$ | $4.002$ ( $3.692$ to $4.292$ ) | 0.152 |
| Participant-level random-effect SDs | Initial severity ($\alpha$; logit scale) | $0.943$ ( $0.753$ to $1.168$ ) | 0.106 |
| Participant-level random-effect SDs | Proportional recovery coefficient ($r$; logit scale) | $0.581$ ( $0.068$ to $1.217$ ) | 0.297 |
| Participant-level random-effect SDs | Time constant ($\tau$; log scale) | $0.782$ ( $0.586$ to $1.015$ ) | 0.109 |

Posterior dependence varied across participants, indicating that parameter trade-offs were not uniform across individuals (Supplementary Figure S2). All 200 parameter-recovery simulations yielded completed fits; 150 (75%) met all final diagnostic criteria and were included in the primary recovery summaries. The remaining 50 failed the R-hat and/or effective-sample-size criteria after one diagnostic-triggered refit (Supplementary Table S1, Panel B). Recovery performance was therefore conditional on the diagnostic-passing fits. These simulations showed differences in individual-level recoverability: median within-simulation Spearman correlations between data-generating values and posterior medians were 0.86 for *α*, 0.33 for *r*, and 0.76 for *τ*, with corresponding calibration slopes of 0.77, 0.15, and 0.60; empirical 95% CrI coverage was 0.92–0.96. Thus, *α* was relatively well recovered, *τ* showed moderate recovery, and *r* showed limited recovery at the individual level (Supplementary Figure S3; Supplementary Table S1). Population- and individual-level parameter estimates were highly similar under wider priors (Supplementary Table S2). Participant-grouped five-fold cross-validation yielded the numerically highest subject-level joint ELPD for the single-exponential model (−689.19), with a small and uncertain difference from the bi-exponential model (ΔELPD, −0.90; SE, 4.06). No evaluated alternative showed a clear advantage over the single-exponential model according to this criterion. Logistic and Gompertz predictions included one fold each that did not meet the E-BFMI criterion (Supplementary Table S3). Full-data expected trajectories and residuals are shown in Supplementary Figure S4.

### Recovery Dynamics Across LCMM-Derived Trajectory Classes

Observed longitudinal SARA trajectories and class-specific Bayesian nonlinear trajectories are shown in Figure 2A. Posterior summaries of the asymptotic proportional recovery coefficient overlapped across classes (r approximately 0.860, 0.864, and 0.835 for Classes 1–3, respectively; Figure 2B). Model-implied proportional improvement by day 180, r(180), was approximately 0.80, 0.78, and 0.53 (Figure 2C), whereas the estimated recovery time constants were approximately 62, 76, and 184 days, respectively (Figure 2D). Thus, the fitted trajectories differed more clearly in recovery timing than in the estimated asymptotic proportional recovery coefficient.

**Figure 2.**
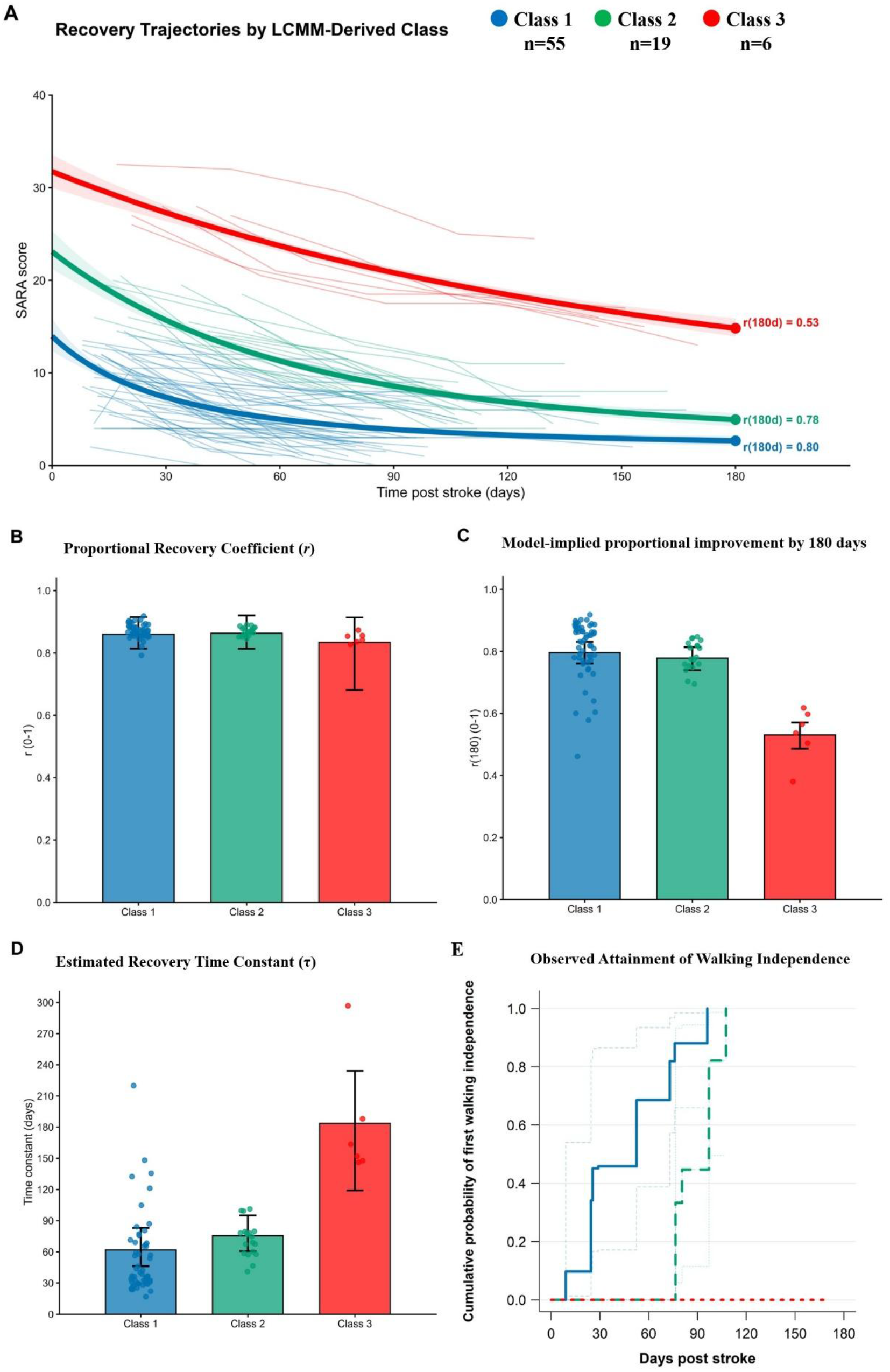
Recovery dynamics and Bayesian nonlinear-model parameters across LCMM-derived trajectory classes. Colors consistently indicate Class 1 (blue), Class 2 (green), and Class 3 (red) across panels. (A) Observed participant-level SARA trajectories and class-specific Bayesian expected trajectories. At each posterior draw and time point, participant-specific expected SARA scores were averaged within each fixed LCMM class. Thick lines show posterior medians of these class-mean trajectories, and shaded areas show pointwise 95% credible intervals (CrIs). Annotations at day 180 show model-implied proportional improvement, r(180). (B–D) Class-specific summaries of (B) the proportional recovery coefficient (r), (C) model-implied proportional improvement by day 180 [r(180)], and (D) the recovery time constant (τ). Points represent participant-specific posterior medians; bars and error bars show posterior medians and 95% CrIs of draw-wise class means. CrIs are conditional on fixed class assignments. Computational details are provided in Supplementary Methods. (E) Turnbull estimates of the cumulative probability of first walking independence, defined as FIM locomotion ≥6 in walking mode. Thin lines indicate 95% confidence intervals for Classes 1 and 2; no first-attainment event was observed in Class 3 during follow-up. SARA, Scale for the Assessment and Rating of Ataxia; LCMM, latent class mixed model; FIM, Functional Independence Measure; CrI, credible interval

### Observed Walking Independence Across Trajectory Classes

Figure 2E shows the cumulative probability of first observed walking independence (FIM locomotion ≥6) from stroke onset. Of the 80 participants, 9 were already independent at the first assessment (left-censored), 61 first attained independence between two assessments (interval-censored), and 10 had not attained independence by the final assessment (right-censored). During the observed follow-up, walking independence was attained by 52 of 55 participants in Class 1 and 18 of 19 in Class 2, whereas none of the 6 participants in Class 3 attained the threshold. The curves showed earlier first attainment in Class 1 than in Class 2, with no observed first-attainment event in Class 3 during follow-up.

### Clinical Characteristics Associated with Model-Derived Recovery Parameters

Posterior-propagated secondary analyses are summarized in Table 3 and Figure 3. In the core model, posterior median contrasts in r were near zero for initial severity, age, sex, and TMT-A, with 95% CrIs including both positive and negative values. Exploratory ROPE probabilities did not support declaring these r contrasts practically equivalent to zero (Supplementary Table S8). For the recovery time constant (τ), poorer TMT-A performance showed a directional association with a longer recovery time scale (median difference, 11.9 days per 1-SD increase in log-transformed completion time; 95% CrI, −2.7 to 32.9). In the exploratory ROPE analysis, 76.6% of the TMT-A–τ posterior contrasts were above the positive ROPE boundary, whereas 22.7% were within the ROPE. In the anatomic sensitivity model, the TMT-A contrast in τ remained positive at the posterior median, although uncertainty persisted after adjustment for stroke subtype and lesion category (9.7 days per 1-SD increase in log-transformed completion time; 95% CrI, −4.0 to 30.4; Table 3). In the functional sensitivity model, higher baseline BBS was associated with a shorter τ, with each 10-point increase corresponding to a median 15.2-day reduction (95% CrI, −27.3 to −5.2); 96.1% of the corresponding posterior contrasts were below the negative ROPE boundary and 3.9% were within the ROPE. The corresponding contrast in r remained uncertain. The α-scale, FAB, and physical therapy sensitivity analyses are reported in Supplementary Table S6.

**Figure 3.**
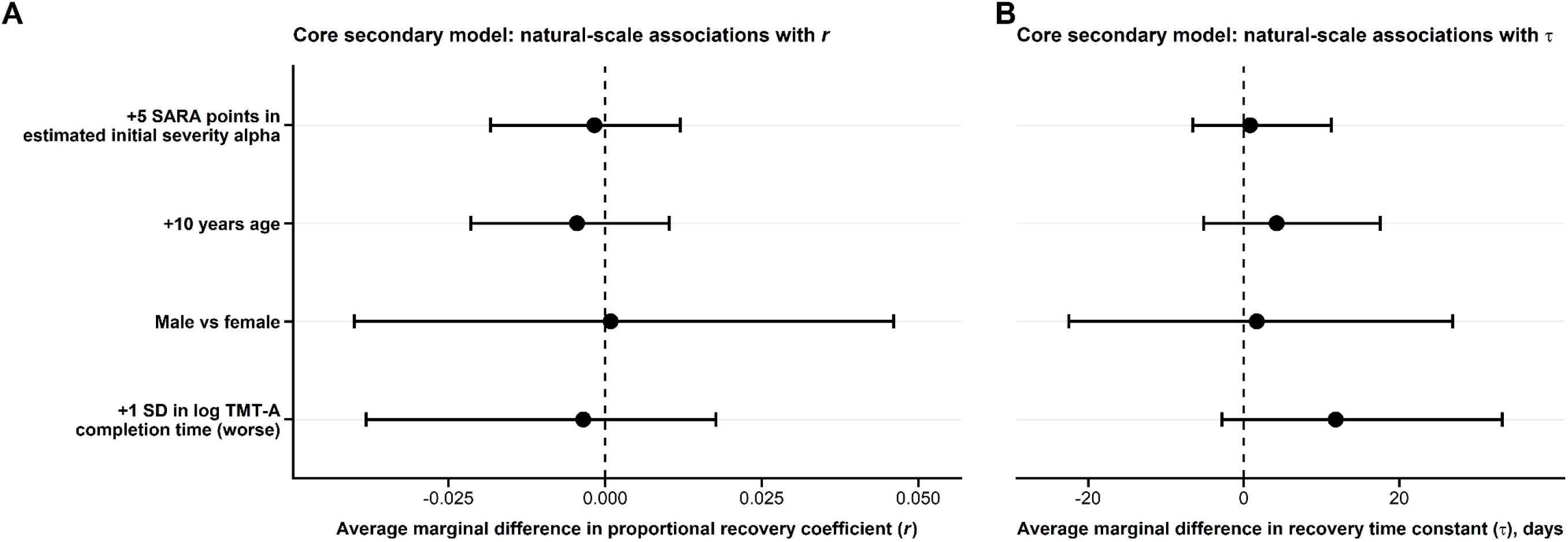
Associations between clinical characteristics and model-derived recovery parameters in the core secondary analysis. Forest plots show posterior-propagated natural-scale associations with (A) the proportional recovery coefficient (r) and (B) the recovery time constant (τ). Points represent posterior medians and horizontal bars represent 95% credible intervals; the dashed vertical line indicates the null value of zero. Estimated initial severity (α) was expressed per 5-point increase on the natural SARA scale, age per 10-year increase, sex as male versus female, and TMT-A per 1-SD increase in log-transformed completion time, with higher TMT-A values indicating poorer performance. Effects for r are expressed as average marginal differences in the proportional recovery coefficient, and effects for τ as average marginal differences in days. Credible intervals incorporate uncertainty in both the participant-level trajectory parameters and the conditional secondary-regression coefficients.

**Table 3.**
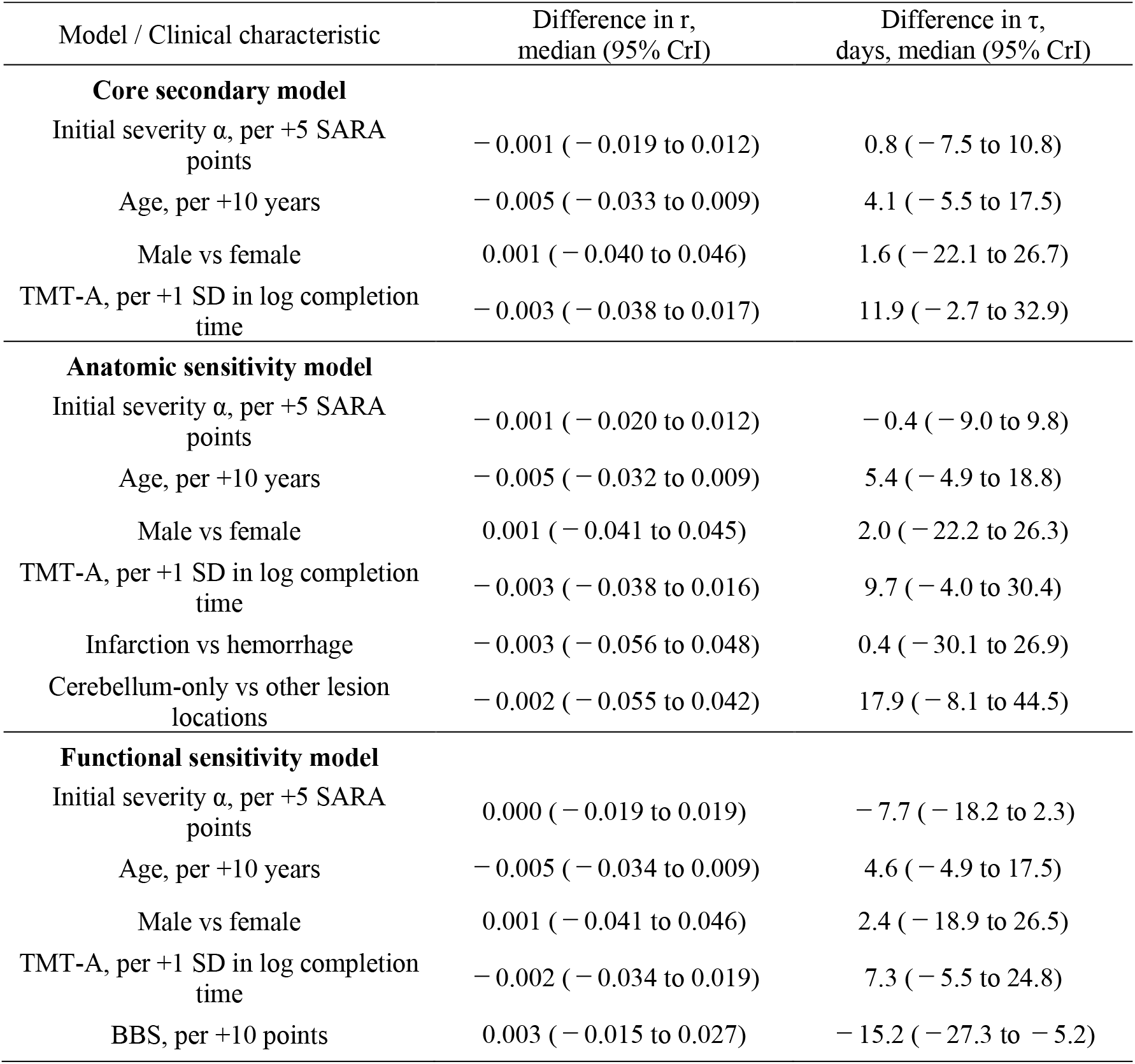
Posterior-propagated secondary analyses of clinical characteristics associated with the proportional recovery coefficient (r) and recovery time constant (τ) Abbreviations: BBS, Berg Balance Scale; CrI, credible interval; SARA, Scale for the Assessment and Rating of Ataxia; TMT-A, Trail Making Test Part A. All models included 80 participants. Values are posterior medians and 95% credible intervals incorporating uncertainty in both the participant-level trajectory parameters and the conditional secondary-regression coefficients. Natural-scale contrasts were calculated as described in the footnote to Supplementary Table S6. Estimated initial severity (α) was entered on its natural SARA scale and expressed per 5-point increase. TMT-A completion time was log-transformed and standardized; a 1-SD increase in log-transformed completion time represents poorer performance. The anatomic sensitivity model additionally included stroke subtype and lesion location, with lesion location categorized as cerebellum-only versus other locations. The functional sensitivity model additionally included baseline BBS. Differences in r are reported on its natural 0-to-1 scale, whereas differences in τ are reported in days. LCMM-derived class membership was not included as a predictor.

## Discussion

Three LCMM-derived classes of post-stroke cerebellar ataxia were characterized using Bayesian nonlinear recovery parameters. Despite broadly overlapping asymptotic proportional recovery (r), the classes differed in estimated initial severity (α) and recovery timing (τ and r(180)). Class 2 had intermediate initial severity but a recovery time scale broadly comparable to Class 1; Class 3 had markedly prolonged τ and lower model-implied proportional improvement by 180 days. Walking independence was observed earlier in Class 1 than in Class 2 and was not observed in Class 3 during follow-up. These findings highlight the importance of considering recovery timing alongside initial severity and proportional improvement.

### Clinical meaning of trajectory classes

Class 2 illustrates the limitations of interpreting post-stroke ataxia recovery solely in terms of initial severity. Previous studies have associated admission SARA scores with concurrent walking status, but the subsequent recovery course remains unclear.^23^ Despite greater admission severity than Class 1, Class 2 achieved most of its model-implied asymptotic recovery by 180 days, and walking independence was observed in nearly all participants during follow-up. These findings suggest that greater initial severity need not imply proportionally less recovery or a markedly prolonged course. Cerebellar reserve, conceptualized as residual adaptive potential within distributed cerebellar networks, may provide a framework for understanding why greater initial impairment does not necessarily preclude improvement.^41,42^ However, this interpretation remains hypothesis-generating, not direct evidence of an underlying recovery mechanism. In contrast, Class 3 showed broadly overlapping asymptotic r but markedly longer τ, lower model-implied proportional improvement by 180 days, and no observed walking independence during follow-up. Initial severity, proportional improvement, recovery timing, and activity-level independence therefore represent related but noninterchangeable dimensions.

### Parameter-level interpretation of recovery

The *α*, *r*, and *τ* parameters decompose the SARA recovery trajectory into initial severity, asymptotic proportional improvement, and the time scale over which recovery becomes clinically expressed. Posterior overlap in r does not establish equivalent biological recovery capacity, particularly given its limited individual-level practical recoverability. Lesion-network mapping has linked anatomically diverse stroke lesions associated with limb ataxia to a shared network involving the cerebellum and cerebellar peduncles.^43^ In a prospective study of isolated cerebellar stroke, the course of ataxia improvement differed by lesion territory.^12^ These findings motivate the hypothesis that lesion location and network involvement may contribute to heterogeneous clinical recovery trajectories, although this possibility was not directly tested in the present study. The early SARA reduction in Figure 2A may partly reflect improvements in gait and postural control. Gait, stance, and sitting contribute substantially to the SARA score,^22^ and balance dysfunction may influence cerebellar gait impairment.^1,2,44^ Improvement in SARA scores may reflect both neurological restitution and compensatory changes in task performance. For example, experimentally modified visual feedback reduced dysmetria during a single-joint reaching task in patients with cerebellar ataxia.^45^ This task-specific effect does not establish the mechanism of longitudinal SARA improvement in the present cohort.

Accordingly, *τ* is more appropriately interpreted as a clinically expressed recovery time scale than as a direct measure of motor-learning speed. Higher baseline BBS was associated with a shorter *τ*. An exploratory ROPE analysis further showed that most of the BBS–*τ* posterior distribution lay beyond the ROPE in the direction of a shorter *τ*, whereas the TMT-A–*τ* distribution was predominantly in the direction of a longer *τ* but remained less definitive with respect to magnitude (Supplementary Table S8). These distribution-based ROPEs were used as relative-magnitude sensitivity thresholds rather than validated clinical-equivalence criteria. The association between baseline balance function and *τ* is observational and may partly reflect overlap between the clinical constructs assessed by the BBS and SARA. Poorer TMT-A performance was directionally associated with a longer *τ*, although uncertainty was substantial. Rather than indicating an attention-specific mechanism, this pattern may reflect broader cognitive-motor resources relevant to adaptation.^21,46–48^ The cerebellar cognitive affective syndrome framework further raises the possibility that non-motor cerebellar dysfunction may contribute to heterogeneity in recovery timing,^20^ although this was not directly assessed. These results characterize the clinical expression and timing of improvement without isolating neurological restitution, compensation, or treatment-specific effects.

### Study Limitations

Several limitations define the level at which these findings should be interpreted. First, the cohort was modest (N=80), and Class 3 included only six participants. Although patient-level bootstrap analyses supported internal stability of class assignment, they do not establish precise class-specific estimates or external reproducibility. The LCMM-derived classes should therefore be regarded as descriptive trajectory patterns that may approximate continuous heterogeneity, rather than established biological subgroups. Second, individual r showed limited practical recoverability, and some τ estimates remained imprecise. Recovery performance was evaluated only in the 150 of 200 simulations that met the diagnostic criteria; exclusion of the remaining 25% limits generalization of these performance estimates to all generated datasets.

In addition, α represents model-estimated severity at stroke onset and therefore depends on extrapolation beyond the earliest observed assessments. The single-exponential model should likewise be regarded as a parsimonious representation of the observed trajectories rather than a unique biological recovery process. These considerations support greater confidence in population- and class-level characterization than in patient-specific biological interpretation. Third, the neuroanatomical and cognitive-motor mechanisms discussed here remain hypothesis-generating because detailed lesion/network analyses and comprehensive assessment of cerebellar cognitive-affective dysfunction were not available. Exclusion of participants with MMSE-J ≤23 further restricted the cognitive range of the cohort, and TMT-A completion time reflects visuomotor as well as attentional performance. Accordingly, associations with BBS and TMT-A should be interpreted as observational clinical associations rather than causal determinants of recovery. Requiring at least three assessments may have selected participants with longer observable rehabilitation courses. Follow-up ended at the last available clinical assessment, and recovery-related differences in discharge or assessment timing may have influenced the observed trajectories. For participants whose final assessment preceded day 180, r(180) involves extrapolation under the fitted model. Furthermore, trajectory classes were identified using the full longitudinal SARA series; their association with observed walking independence should therefore not be interpreted as validation of a prognostic classification available at admission.

## Conclusions

Three LCMM-derived trajectory classes exhibited heterogeneous patterns of recovery from post-stroke cerebellar ataxia, distinguished by model-estimated initial severity (*α*) and particularly recovery time scale (*τ*), despite broadly overlapping asymptotic proportional recovery (*r*). Characterizing both improvement magnitude and timing may be more clinically informative than initial severity alone. Validation in larger independent cohorts is required before patient-level prognostic application.

## Data Availability

All data produced in the present study are available upon reasonable request to the authors

## Acknowledgments

We received valuable advice and assistance from the staff of the Rehabilitation Department at the Maruki Memorial Medical and Social Welfare Center, a Social Welfare Corporation, and the staff of the Neurorehabilitation Laboratory, Graduate School of Health Sciences, Kio University.

## Author contributions

**Yuichiro Yamasaki**: Conceptualization; Data curation; Formal analysis; Investigation; Funding acquisition; Resources; Methodology; Project administration; Writing - original draft; Writing - review & editing.

**Yusaku Takamura**: Conceptualization; Formal analysis; Methodology; Writing - review & editing.

**Hirofumi Sato**: Data curation; Investigation; Methodology; Project administration.

**Katsunobu Okuma**: Data curation; Investigation; Methodology; Project administration.

**Yohei Kobayashi**: Data curation; Investigation; Methodology; Project administration.

**Ariyasu Kamijima**: Data curation; Investigation; Methodology.

**Shinjiro Takaishi**: Data curation; Investigation; Methodology.

**Hideyuki Maruki**: Project administration.

**Shu Morioka**: Conceptualization; Methodology; Funding acquisition; Supervision; Writing - review & editing.

## Ethical Considerations

This study was conducted with the approval of the ethics committees at Maruki Memorial Medical and Social Welfare Center, Saitama Citizens Medical Center, and Saitama Sekishinkai Hospital (approval numbers: Maruki Memorial Medical and Social Welfare Center: 32; Saitama Citizens Medical Center: 2020-09; Saitama Sekishinkai Hospital: 2023-5). All participants provided written informed consent in accordance with the Declaration of Helsinki.

## Data Availability

Data supporting the findings of this study are available from the corresponding author upon request. The analysis code has not been deposited in a public repository.

