## Supplemental Tables and Figures for "Recovery Trajectories in Post-stroke Ataxia: Distinct Longitudinal Classes Characterized by Initial Severity and Recovery Timing"

Neurorehabilitation and Neural Repair

Manuscript ID: NNR-26-0187

This supplement contains additional methodological details, six supplementary figures, and eight supplementary tables.

### Contents

**Supplementary Methods**

1. Parameter-recovery simulations……………………………………………………2
2. Prior-sensitivity analysis………………………………………………………… 2
3. Alternative-model comparison and cross-validation……………………………. 3
4. LCMM estimation and class-specific summaries………………………………...4
5. Exploratory ROPE analysis of posterior-propagated associations……………….6
6. Supplementary Figures S1–S6…………………………………………………....7
7. Supplementary Tables S1–S8……………………………………………………16

### Supplementary Methods

#### 1. Parameter-recovery simulations

Parameter-recovery simulations reproduced the study's 80-participant, 283-observation assessment schedule. For each simulated dataset, one joint posterior draw was selected without replacement from the 8,000 post-warm-up draws of the fitted primary model. Participant-specific trajectory parameters and the residual standard deviation were taken from the same draw. Gaussian observations were generated at the original assessment times, retaining the participant-specific effects from that draw. Simulated observations were neither rounded nor truncated to the SARA range of 0–40. Each simulated dataset was refitted using the primary model's likelihood, hierarchical structure, and original priors. Initial fits used four chains of 4,000 iterations, including 2,000 warm-up iterations, with adapt_delta = 0.99 and max_treedepth = 15. Fits not meeting the diagnostic criteria were refitted once using 6,000 iterations per chain, including 3,000 warm-up iterations, with adapt_delta = 0.995 and max_treedepth = 15. Acceptance required a maximum R-hat ≤1.01, minimum bulk and tail effective sample sizes ≥400, no divergent transitions or maximum-treedepth hits, and a minimum chain-specific E-BFMI ≥0.30. Primary recovery summaries included only final fits meeting all these criteria. Recovery metrics compared the data-generating values with recovered posterior medians; the reported performance therefore pertains to this fitted-model simulation setting and diagnostic-passing analysis set. Simulation completion, refitting, and final diagnostic outcomes are summarized in Supplementary Table S1, Panel B.

#### 2. Prior-sensitivity analysis

The sensitivity specification retained the primary model's likelihood and hierarchical structure while doubling the standard deviations of the Normal priors and the scale parameters of the half-Cauchy priors, without changing their location parameters. The population-level intercepts for $a_i$ and $p_i$ were assigned Normal (0, 4) priors, and the intercept for $\lambda_i$ was assigned a Normal (3, 2) prior; the second argument denotes the standard deviation. The corresponding random-effect standard deviations were assigned half-Cauchy priors with scale parameters 4, 2, and 2, respectively, and the residual standard deviation was assigned a half-Cauchy prior with scale parameter 10. Comparisons included natural-scale population-level summaries obtained by transforming the population intercepts conditional on zero random effects, participant-specific posterior medians, and random-effect and residual standard deviations. Results are reported in Supplementary Table S2.

### 3. Alternative-model comparison and cross-validation

All candidate models used a Gaussian likelihood for SARA scores on their original scale. The same participant-grouped five-fold partition was used across models, with each participant's complete longitudinal series assigned to a single fold. Participants were ordered by first-assessment SARA, follow-up duration, and identifier; within consecutive blocks of five, one participant was randomly allocated to each fold. At each fold, models were fitted to 64 participants (80% of the cohort) and evaluated on the complete longitudinal series of the remaining 16 participants (20%). All observations from a participant were kept in the same fold, so no participant contributed observations to both the training and test sets within a fold. Each participant was held out once across the five folds. These percentages refer to participants, not individual observations. Time-standardization constants and the linear/spline intercept priors used full-cohort information, as detailed in Supplementary Table S3, and were held fixed across folds rather than re-estimated using training data alone. Predictions for held-out participants used 2,000 draws, with new participant-level random effects sampled from the Gaussian distribution implied by each training fit's posterior group-level parameters. Within each draw, the same participant-specific effects were shared across that participant's assessment times. The participant-level joint log predictive density was calculated by summing observation-level log likelihoods within each draw and then taking the log of the mean of the exponentiated sums across draws. Total ELPD was the sum over the 80 held-out participants. Standard errors of between-model ELPD differences were calculated from paired participant-level differences as $\sqrt{[N \times \mathrm{Var}(d_i)]}$, where $N = 80$ and $d_i$ denotes the participant-specific difference. RMSE was calculated from the posterior median of the expected SARA score at each held-out observation. The 95% predictive intervals were defined by the 2.5th and 97.5th percentiles of posterior predictive draws, including residual variation. RMSE and predictive interval coverage were pooled across all 283 held-out observations rather than averaged across participant-level summaries. Predictions from all five folds were retained for each model, with diagnostic status reported separately in Supplementary Table S3.

### 4. LCMM estimation and class-specific summaries

#### Model specification and class enumeration

Linear random-intercept and quadratic common-curvature random-intercept models were initially evaluated with one to three classes. Both structures were subsequently extended to four classes as an additional sensitivity analysis, retaining the same analysis data, outcome standardization, time transformation, and random-effect and residual-variance structures. The expanded candidate set therefore comprised eight models. SARA scores were standardized using the mean and standard deviation of all longitudinal observations. Time was defined as (days since stroke onset − median first-assessment day across participants)/30. In multiple-class models, intercepts and linear time slopes were class-specific, whereas the random-intercept variance and residual variance were common across classes. The quadratic coefficient was also common across classes in the quadratic models. Each multiple-class candidate was fitted using a direct initialization from the one-class model and four independent batches of 25 random starts. Optimization reproducibility required at least two converged solutions within 0.05 log-likelihood units of the best solution and a minimum pairwise adjusted Rand index (ARI) of 0.95 among these solutions. Absolute and preferred minimum class sizes were five and eight participants, respectively; these were internal assessment criteria rather than universal thresholds. BIC was the primary class-enumeration criterion; AIC, SABIC, ICL1/ICL2, classification quality, class size, and numerical reproducibility were examined as complementary information. The four-class models were not included in the participant-level bootstrap assessment of the retained three-class model. For multiple-class solutions, the preferred flag shown in Figure 1A required numerical admissibility, a minimum class size of eight, entropy ≥0.70, and mean maximum posterior membership probability ≥0.70. This flag was a supplementary assessment and did not determine the final class-number decision.

### Bootstrap resampling and optimization

Bootstrap samples were generated by sampling participants with replacement and retaining each sampled participant's longitudinal observations. The selected three-class model structure was held fixed. Each fitted model was used to classify all 80 original participants. Class labels were aligned to the full-data reference trajectories by minimizing the total mean squared trajectory difference across matched classes. Stage 1 compared a reference-model-initialized benchmark, a direct fit initialized from a bootstrap-specific one-class model, and a 10-start grid search. Stage 2 added five batches of 10 random starts when triggered by prior initialization-stress-test status, optimization failure, a log-likelihood gain greater than 0.10 over the benchmark, partition disagreement with the benchmark (ARI <0.90 or smallest-class Jaccard <0.95), near-best partition disagreement (ARI <0.95), or a smallest class containing fewer than five original participants. Stage 3 added another five batches of 10 starts when optimization remained unresolved. The final candidate was selected by the highest observed finite log-likelihood among converged solutions, not by agreement with the reference partition. A replicate was labeled optimization-resolved if no escalation criterion was met at Stage 1 or, after escalation, at least two solutions were within 0.01 log-likelihood units of the best solution and their minimum pairwise ARI was at least 0.95. A usable fitted solution and full-cohort classification could therefore be available even when optimization remained unresolved. All successful replicates were retained in the stability summaries regardless of optimization-resolution status. ARI compared the full-cohort partitions; class-specific Jaccard indices compared membership sets. Participant-level same-class frequency was the proportion of successful replicates assigning that participant to the reference class.

### Class-specific Bayesian summaries

Class membership was held fixed at the maximum-posterior-probability assignment. Within each joint Bayesian posterior draw, participant-specific α, r, and τ were transformed to their natural scales, and $r_i(180)$ was calculated before averaging within each class. Parameter summaries used all 8,000 post-warm-up draws. Class-specific expected trajectories were calculated using 2,000 approximately evenly spaced draws by averaging participant-specific expected SARA scores within each class at each time point, rather than evaluating a trajectory at class-average parameters. Reported summaries were the posterior median and the 2.5th and 97.5th percentiles of these draw-wise class means. Trajectory intervals were pointwise credible intervals for the class-mean expected trajectory, not individual prediction intervals. These intervals represent Bayesian parameter uncertainty conditional on fixed class assignments and do not propagate LCMM class-assignment uncertainty. Membership uncertainty was summarized separately in Supplementary Table S5.

### 5. Exploratory ROPE analysis of posterior-propagated associations

To provide an additional context for the magnitude of the posterior-propagated secondary associations, an exploratory region-of-practical-equivalence (ROPE) analysis was performed on the natural parameter scales. For each primary-model posterior draw, the between-participant standard deviation of the natural-scale r and $\tau$ values was calculated across the 80 participants. The posterior median of these standard deviations was used as a fixed reference, and the exploratory ROPE was defined as ±0.10 of the corresponding reference standard deviation. The exploratory 0.10-SD criterion was informed by commonly used standardized ROPE conventions.[1] This yielded ROPE bounds of ±0.00655 for r and ±6.18 days for $\tau$. For each clinical contrast, the proportions of the 4,000 propagated natural-scale posterior contrasts falling below, within, and above the corresponding ROPE were calculated. The analysis was applied to all contrasts in the core secondary model and to the additional BBS contrast from the functional sensitivity model. The ROPE was used as a distribution-based relative-magnitude sensitivity criterion and was not interpreted as a validated clinical-equivalence threshold. Results are reported in Supplementary Table S8.

### Supplementary References

Makowski D, Ben-Shachar MS, Chen SHA, Lüdecke D. Indices of Effect Existence and

Significance in the Bayesian Framework. Front Psychol. 2019;10:2767. doi:10.3389/fpsyg.2019.02767.

### 6. Supplementary Figures S1–S6.

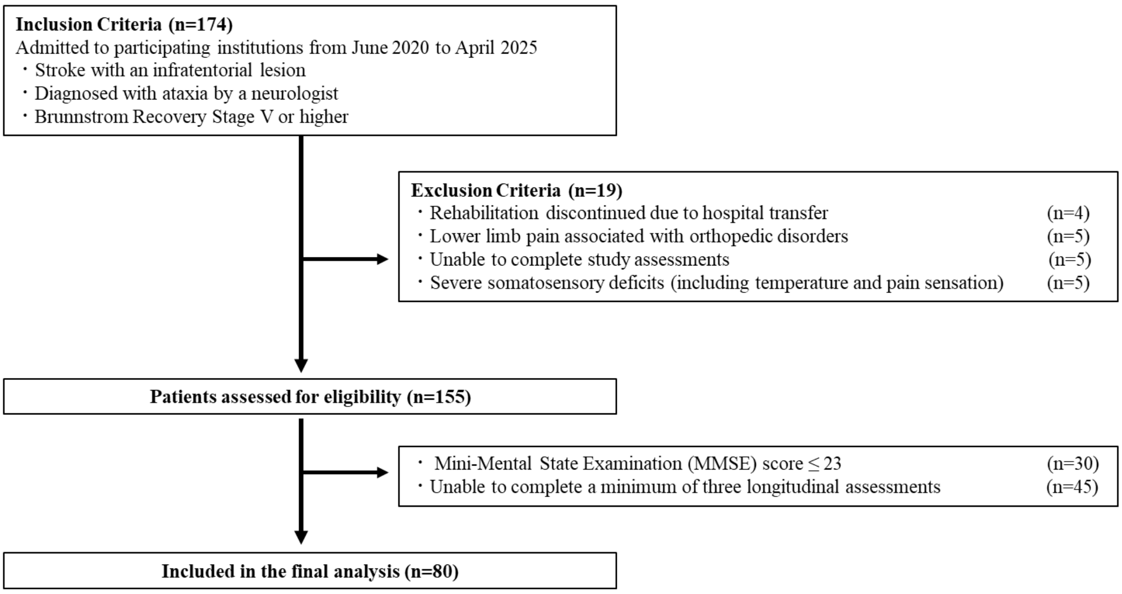


**Supplementary Figure S1. Flowchart of Participant Selection.**

Flowchart showing participant screening, exclusion, and inclusion in the final analysis. Of 174 patients screened, 80 participants meeting the eligibility and longitudinal-assessment criteria were included in the final analysis.

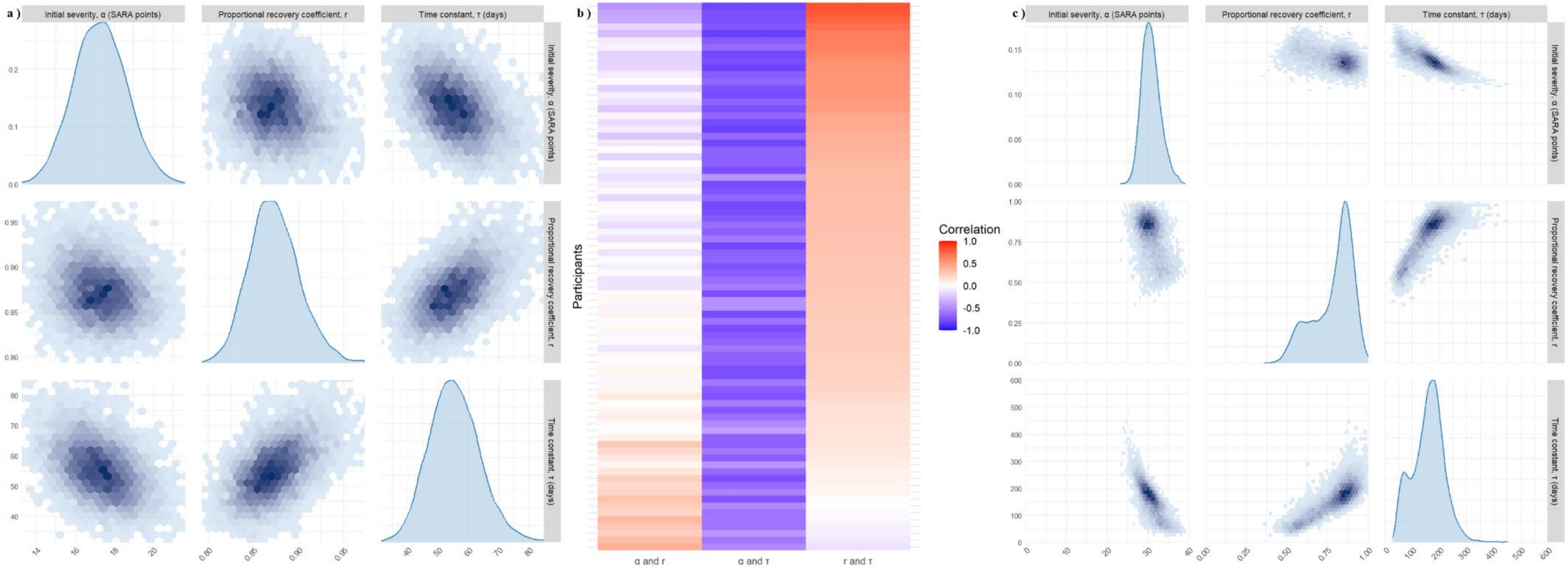


**Supplementary Figure S2. Posterior dependence among population- and individual-level model parameters.**

(A) Marginal and pairwise joint posterior distributions of the population-level initial severity (α, SARA points), proportional recovery coefficient (r), and recovery time constant (τ, days), transformed to their natural scales and evaluated conditional on zero random effects. Axes were restricted to the posterior support to facilitate visualization; the theoretical domains were 0–40 for α, 0–1 for r, and >0 days for τ. (B) Within-participant Pearson correlations across posterior draws for α–r, α–τ, and r–τ in all 80 participants. Rows represent participants, with the same ordering across columns; colors range from −1 to 1. (C) Marginal and pairwise joint posterior distributions for the participant with the largest within-participant r–τ posterior correlation (0.829), selected using a prespecified objective criterion to illustrate the upper end of observed posterior dependence. Darker shading in panels A and C denotes greater posterior density.  SARA, Scale for the Assessment and Rating of Ataxia.

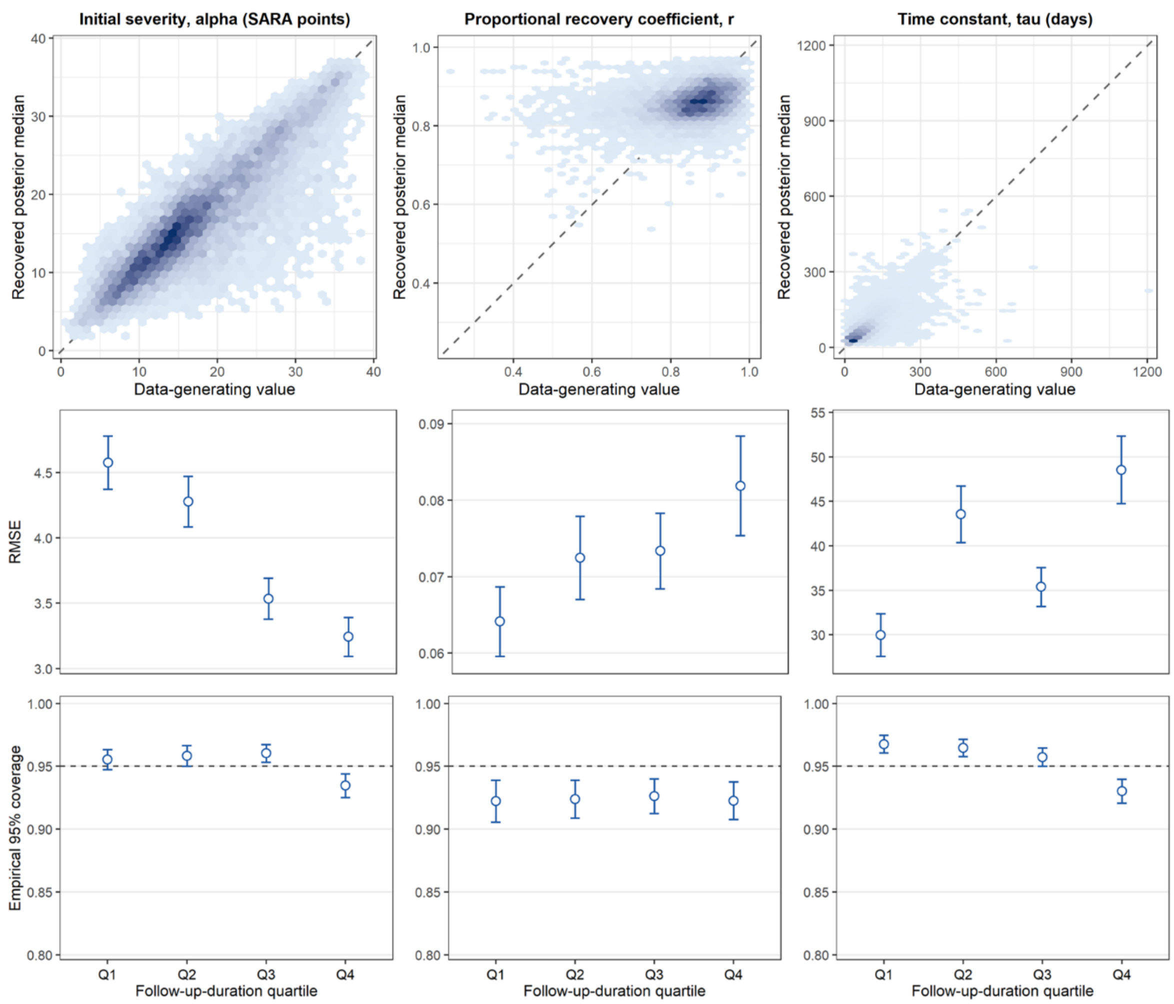


**Supplementary Figure S3. Individual-level parameter recovery under the observed assessment schedule.**

The upper row compares data-generating values with recovered posterior medians for individual α, r, and τ; dashed lines indicate identity. The middle and lower rows show, respectively, root mean squared error (RMSE) and empirical 95% credible-interval (CrI) coverage by quartile of the interval between each participant's first and final SARA assessments. Results are based on the 150 of 200 completed simulations meeting the diagnostic criteria described in Supplementary Methods, "Parameter-recovery simulations"; each simulated dataset followed the study's 80-participant, 283-observation assessment schedule. Points in the middle and lower rows show mean performance across simulations, and error bars show 95% Monte Carlo confidence intervals. The horizontal dashed line in the coverage panels denotes the nominal 0.95 level. Follow-up-duration quartiles were Q1, 39–60 days (n=20); Q2, 60–69 days (n=20); Q3, 69–90 days (n=20); and Q4, 90–148 days (n=20). α, estimated initial severity; r, proportional recovery coefficient; τ, recovery time constant; CrI, credible interval; RMSE, root mean squared error; SARA, Scale for the Assessment and Rating of Ataxia.

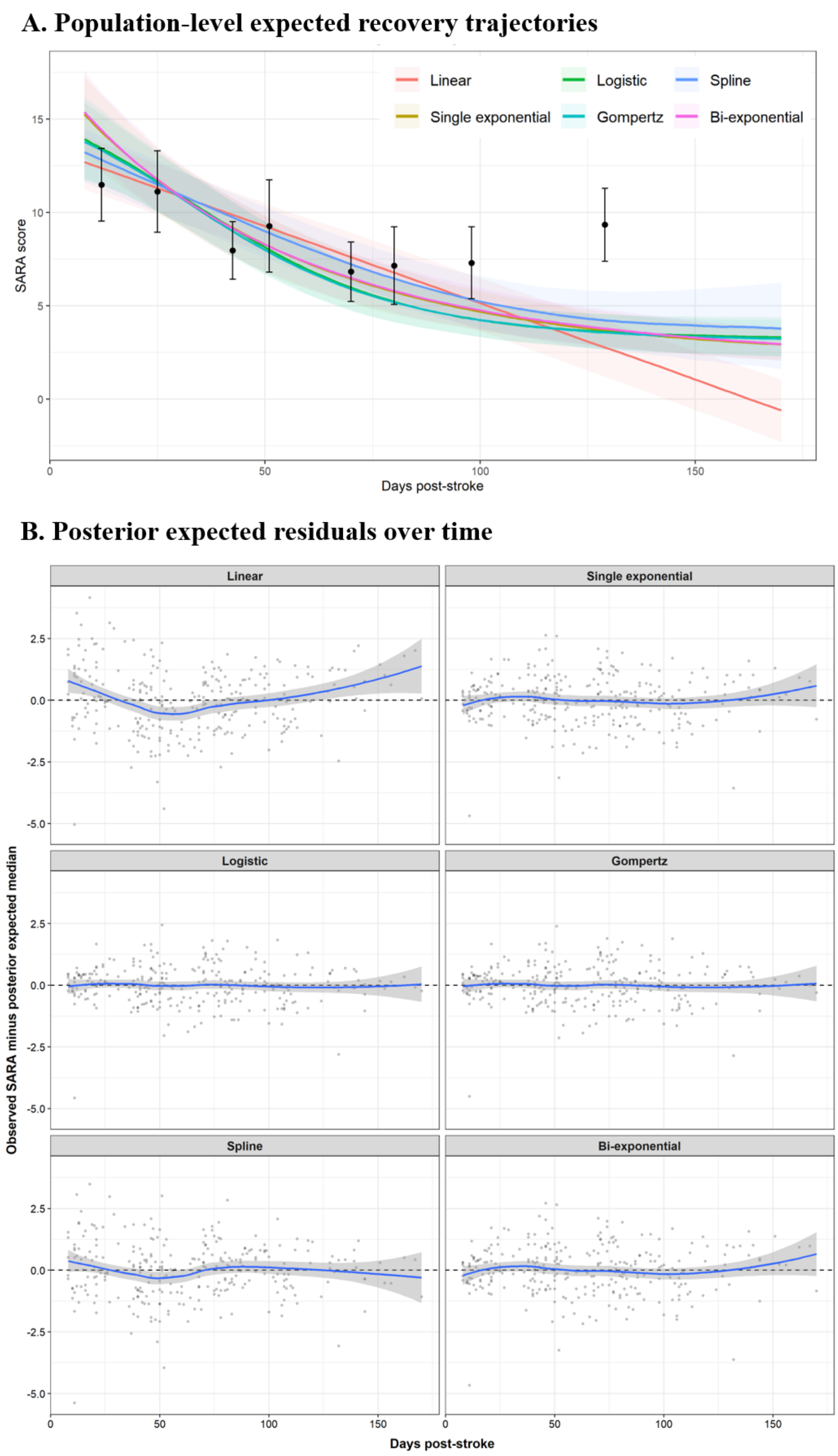


**Supplementary Figure S4. Posterior predictive assessment of candidate longitudinal recovery models.**

(A) Expected SARA trajectories from the full-data fits of the six candidate models, evaluated conditional on zero participant-specific random effects. Colored lines indicate posterior medians, and shaded bands indicate pointwise 95% credible intervals for the expected trajectories. Black points indicate observed mean SARA scores within quantile-based time bins, with error bars representing mean ±1.96 SE. (B) Observation-level residuals from the full-data fits, defined as observed SARA minus the posterior median of the participant-specific expected SARA score, including estimated participant-level random effects, plotted against days post-stroke. Gray points indicate individual observations, blue lines indicate LOESS smooths, shaded bands indicate 95% confidence intervals for the smooths, and dashed horizontal lines indicate zero residual. Both panels describe fit to the observed cohort rather than the held-out predictions used in Supplementary Table S3. CrI, credible interval; LOESS, locally estimated scatterplot smoothing; SARA, Scale for the Assessment and Rating of Ataxia; SE, standard error.

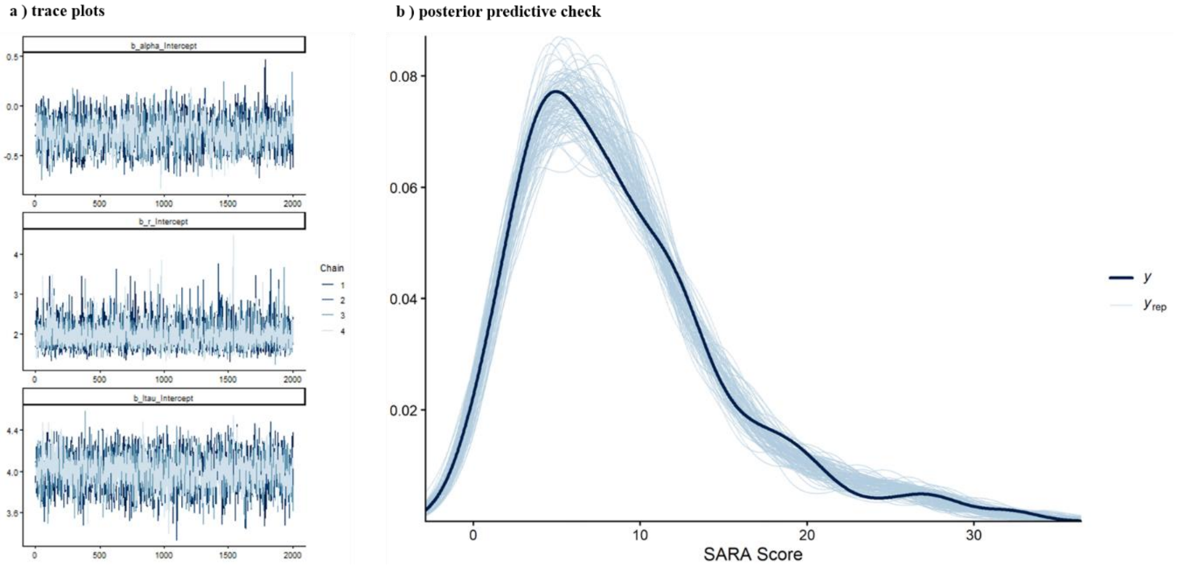


**Supplementary Figure S5. MCMC diagnostics and posterior predictive check for the primary Bayesian nonlinear mixed-effects model.**

(A) Trace plots for the population-level intercepts of the internal predictors $a_i = \text{logit}(\alpha_i/40)$, $p_i = \text{logit}(r_i)$, and $\lambda_i = \log(\tau_i - 0.142)$ across four MCMC chains after warm-up. (B) Posterior predictive density check for the observed SARA scores. The dark line represents the observed-data density (y), and the light lines represent replicated datasets (y_rep) generated from the posterior predictive distribution. MCMC, Markov chain Monte Carlo; SARA, Scale for the Assessment and Rating of Ataxia.

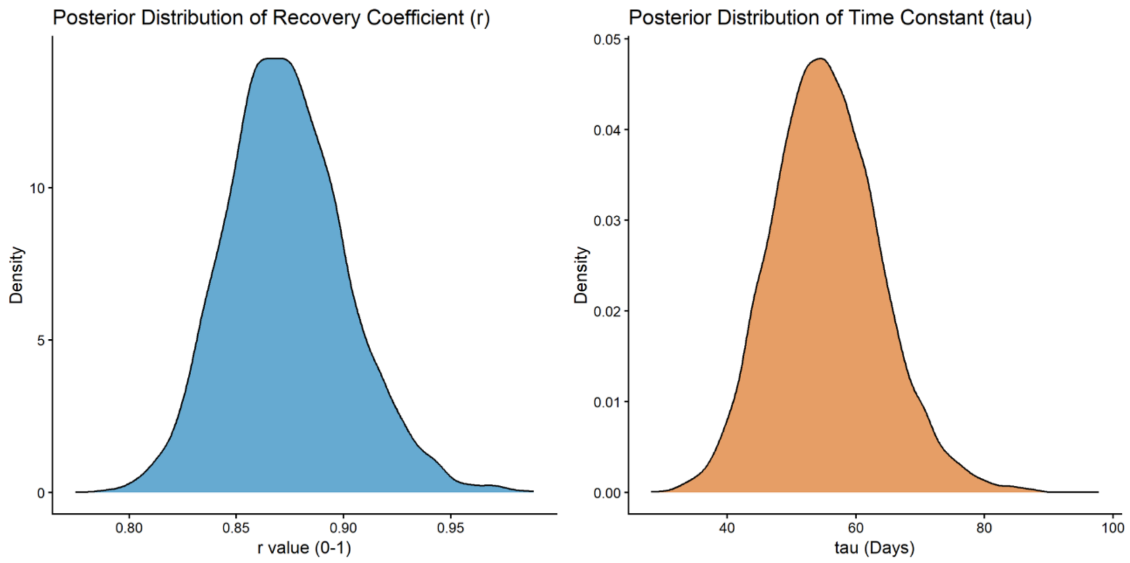


**Supplementary Figure S6. Natural-scale posterior distributions of r and τ.**

Posterior density distributions of the population-level $r$ and τ parameters after transformation from their model-internal intercept scales to the natural scales, evaluated conditional on zero random effects. The proportional recovery coefficient was transformed as $r = \text{logistic}(\eta_r)$, and the recovery time constant as $\tau = \exp(\eta_\tau) + 0.142\text{days}$. τ represents the characteristic time scale of the exponential recovery function: at $t = \tau$, approximately 63.2% $(1 - e^{-1})$of the model-implied recoverable change has occurred. Larger τ values therefore indicate a slower approach toward the model-implied asymptote.

### 7. Supplementary Tables S1–S8.

**Supplementary Table S1. Parameter-recovery performance and simulation diagnostics.**

**Panel A. Parameter-recovery performance in 150 diagnostic-passing fits.**

| Level | Parameter | Bias | RMSE | 95% CrI coverage | CrI width[a] | Median within-Simulation Spearman ρ | Calibration slope |
|---|---|---|---|---|---|---|---|
| Population | α | −0.12 | 1.38 | 0.95 | 5.68 | — | 0.29 |
| Population | r | −0.01 | 0.03 | 0.93 | 0.11 | — | 0.56 |
| Population | τ | −0.14 | 10.32 | 0.95 | 38.3 | — | 0.26 |
| Individual | α | −0.75 | 4.09 | 0.95 | 15.0 | 0.86 | 0.77 |
| Individual | r | 0.00 | 0.08 | 0.92 | 0.27 | 0.33 | 0.15 |
| Individual | τ | −4.21 | 44.00 | 0.96 | 148.8 | 0.76 | 0.60 |

**Panel B. Simulation completion, refitting, and final diagnostic outcomes.**

| Diagnostic outcome | n/N (%) |
|---|---|
| Completed simulated datasets | 200/200 (100%) |
| Met all criteria at the initial fit | 113/200 (56.5%) |
| Underwent one diagnostic-triggered refit | 87/200 (43.5%) |
| Met all criteria after refitting, among refitted datasets | 37/87 (42.5%) |
| Final fits meeting all criteria | 150/200 (75.0%) |
| Final fits not meeting all criteria | 50/200 (25.0%) |
| R-hat criterion only | 6/50 (12.0%) |
| One or both ESS criteria only | 19/50 (38.0%) |
| Both R-hat and ESS criteria | 25/50 (50.0%) |

Bias was calculated as the recovered posterior median minus the data-generating value. RMSE = root mean squared error; CrI = credible interval. Coverage is the proportion of 95% CrIs containing the data-generating value. [a] CrI width is the arithmetic mean width of the 95% credible intervals across the included estimates at each level. Within-simulation Spearman ρ was calculated across the 80 participants in each simulation and summarized by the median across simulations. Calibration slopes were obtained by regressing recovered posterior medians on data-generating values after pooling the estimate–truth pairs across retained simulations, separately for each parameter and estimation level. Panel A reports recovery performance conditional on the 150 final fits satisfying all diagnostic criteria. Panel B summarizes completion, refitting, and diagnostic outcomes. The three failure categories are mutually exclusive: an R-hat failure denotes a maximum R-hat >1.01, and an ESS failure denotes a minimum bulk and/or tail ESS <400. Across all 200 final fits, there were no divergent transitions or maximum-treedepth hits, and the minimum chain-specific E-BFMI was 0.434. Diagnostic acceptance does not itself establish accurate parameter recovery, and the Panel A estimates do not describe unconditional performance across all 200 simulated datasets. Data-generating-process alignment was verified in 5 of 5 sampled draws, with a maximum absolute difference <0.001.

**Supplementary Table S2. Prior sensitivity of population-level, individual-level, and variance-component estimates.**

**A. Population-level parameters.**

| Parameter | Original-prior model, median (95% CrI) | Wider-prior model, median (95% CrI) | Median difference[a] |
|---|---|---|---|
| α, SARA points | 17.1 (14.6–19.9) | 17.1 (14.6–19.9) | −0.09 |
| r | 0.87 (0.82–0.93) | 0.88 (0.83–0.95) | +0.006 |
| τ, days | 55.1 (40.4–73.1) | 56.6 (41.2–76.7) | +1.44 days |

**B. Individual-level agreement.**

| Parameter | Pearson r | Spearman ρ | Mean absolute difference | Maximum absolute difference |
|---|---|---|---|---|
| α | 0.99 | 0.99 | 0.12 SARA points | 0.57 SARA points |
| r | 0.99 | 0.99 | 0.006 | 0.011 |
| τ | 0.99 | 0.99 | 1.35 days | 5.04 days |

**C. Group-level and residual standard deviations.**

| Variance component | Original-prior model, median (95% CrI) | Wider-prior model, median (95% CrI) | Relative median change[b] |
|---|---|---|---|
| Random-intercept SD: logit(α/40) | 0.94 (0.76–1.16) | 0.94 (0.76–1.18) | +0.1% |
| Random-intercept SD: logit(r) | 0.56 (0.07–1.23) | 0.65 (0.07–1.40) | +15.4% |
| Random-intercept SD: log(τ − 0.142) | 0.78 (0.58–1.02) | 0.77 (0.59–1.01) | −0.9% |
| Residual SD | 1.34 (1.18–1.52) | 1.34 (1.17–1.52) | −0.2% |

The original and wider prior specifications and the quantities compared are described in Supplementary Methods, Prior-sensitivity analysis. CrI = credible interval. [a] Median difference was calculated as the wider-prior posterior median minus the original-prior posterior median. Pearson r and Spearman ρ compare individual posterior medians under the two prior specifications. [b] Relative median change was calculated as 100 × (wider-prior median − original-prior median)/original-prior median. Random-effect standard deviations are reported on the logit(α/40), logit(r), and log(τ − 0.142) scales, respectively; the residual SD is reported in SARA points.

**Supplementary Table S3. Predictive comparison of alternative longitudinal recovery models.**

| Model | Full-data MCMC | K-fold diagnostic criteria met, n/5 | Subject-level joint ELPD | ΔELPD vs exponential | SE of ΔELPD | RMSE, SARA points | 95% predictive interval coverage |
|---|---|---|---|---|---|---|---|
| Single exponential | Pass | 5/5 | −689.19 | 0 | — | 6.53 | 0.908 |
| Bi-exponential | Pass | 5/5 | −690.09 | −0.90 | 4.06 | 6.51 | 0.901 |
| Logistic | Pass | 4/5* | −695.00 | −5.81 | 5.80 | 6.47 | 0.887 |
| Gompertz | Pass | 4/5* | −695.17 | −5.97 | 4.58 | 6.49 | 0.901 |
| Spline | Pass | 5/5 | −720.70 | −31.50 | 8.43 | 6.34 | 0.926 |
| Linear | Pass | 5/5 | −736.21 | −47.02 | 15.46 | 6.60 | 0.908 |

Full-data MCMC diagnostics refer to models fitted to all observations. Predictive performance measures were obtained from participant-grouped five-fold cross-validation, with 64 training participants (80%) and 16 held-out participants (20%) per fold, as described in Supplementary Methods, "Alternative-model comparison and cross-validation." Time-standardization constants were calculated from the full cohort. For the linear and spline models, the Normal intercept priors were specified using the full-cohort SARA mean and standard deviation and were retained across folds. These data-dependent settings were therefore not estimated exclusively within each training fold. "Pass" indicates that the full-data fit met all prespecified diagnostic criteria: R-hat ≤1.01, bulk effective sample size (ESS) ≥400, tail ESS ≥400, E-BFMI ≥0.30 in all chains, zero divergent transitions, and zero maximum-treedepth hits. K-fold diagnostic status indicates the number of folds meeting the same criteria. *For the logistic and Gompertz models, fold 5 did not meet the E-BFMI criterion (minimum E-BFMI, 0.264 and 0.252, respectively); the other prespecified criteria were met in those folds. Predictions from these folds were retained in the reported performance measures; comparisons involving these models should be interpreted with this diagnostic limitation. ΔELPD was calculated as the model ELPD minus the single-exponential model ELPD. ELPD, expected log predictive density; E-BFMI, energy Bayesian fraction of missing information; ESS, effective sample size; MCMC, Markov chain Monte Carlo; RMSE, root mean square error; SE, standard error.

**Supplementary Table S4. Class-enumeration results for candidate latent class mixed models.**

**Panel A. Penalized fit criteria.**

| Structure | G | p | AIC | BIC | SABIC | ICL1 | ICL2 |
|---|---|---|---|---|---|---|---|
| Linear RI | 1 | 4 | 437.27 | 446.80 | 434.18 | 446.80 | 446.80 |
| Linear RI | 2 | 7 | 421.88 | 438.55 | 416.48 | 466.06 | 467.62 |
| Linear RI | 3 | 10 | 394.91 | 418.73 | 387.20 | 448.82 | 454.57 |
| Linear RI | 4 | 13 | 392.03 | 423.00 | 382.00 | 456.81 | 458.97 |
| Quadratic CC-RI | 1 | 5 | 421.86 | 433.77 | 418.01 | 433.77 | 433.77 |
| Quadratic CC-RI | 2 | 8 | 378.09 | 397.15 | 371.92 | 405.76 | 405.04 |
| Quadratic CC-RI | 3 | 11 | 354.47 | 380.68 | 345.99 | 391.31 | 391.05 |
| Quadratic CC-RI | 4 | 14 | 352.85 | 386.20 | 342.05 | 416.18 | 417.42 |

**Panel B. Classification quality, class sizes, and numerical reproducibility.**

| Structure | G | Entropy | Mean max probability | Class sizes (descending) | Near-best final fits | Minimum ARI |
|---|---|---|---|---|---|---|
| Linear RI | 1 | — | 1.000 | 80 | — | — |
| Linear RI | 2 | 0.504 | 0.847 | 64 / 16 | 5 | 1.000 |
| Linear RI | 3 | 0.658 | 0.818 | 46 / 28 / 6 | 5 | 1.000 |
| Linear RI | 4 | 0.695 | 0.818 | 29 / 25 / 20 / 6 | 5 | 1.000 |
| Quadratic CC-RI | 1 | — | 1.000 | 80 | — | — |
| Quadratic CC-RI | 2 | 0.845 | 0.957 | 63 / 17 | 5 | 1.000 |
| Quadratic CC-RI | 3 | 0.879 | 0.946 | 55 / 19 / 6 | 5 | 1.000 |
| Quadratic CC-RI | 4 | 0.730 | 0.838 | 41 / 18 / 15 / 6 | 5 | 1.000 |

All models used the same 80 participants and 283 observations. Information criteria were calculated from the maximized log-likelihood on the standardized SARA scale; N = 80 participants was used for BIC and SABIC. G = 4 rows denote the additional sensitivity models. BIC was the primary selection criterion. For G = 1, entropy and classification-reproduction statistics are not applicable; the membership probability of 1 is trivial. For each G >1 model, the five final solutions comprise the direct fit and the four grid-search batch winners, not all 100 short-start runs. Numerical reproducibility is distinct from participant-bootstrap reproducibility.

Table S4 reports overall classification entropy as one minus the mean participant-level normalized entropy, whereas Supplementary Table S5, Panel B reports the median participant-level normalized entropy within each class. The three-class quadratic model had the lowest BIC and ICL1/ICL2 among the eight candidates. Increasing the number of classes from three to four increased BIC by 5.52 for the quadratic structure and by 4.26 for the linear structure. The criteria were not unanimous: the four-class quadratic model had lower AIC (352.85 vs 354.47) and SABIC (342.05 vs 345.99).

For each four-class candidate, the direct fit and all four grid-search batch winners converged within 0.05 log-likelihood units of the best solution, with minimum pairwise ARI = 1.000. The quadratic four-class solution comprised 41, 18, 15, and 6 participants in descending size order. The same six participants remained together in the smallest class, while the larger groups were repartitioned.

Retention of the three-class model was based on BIC-led evaluation supported by ICL and clearer classification, not on failure to fit a fourth class or unanimous agreement of all criteria.

BIC, Bayesian information criterion; SABIC, sample-size-adjusted BIC; ICL, integrated completed likelihood; LCMM, latent class mixed model. G, number of latent classes; p, number of freely estimated parameters; RI, random intercept; CC-RI, common-curvature random-intercept; AIC, Akaike information criterion; ARI, adjusted Rand index.

**Supplementary Table S5. Bootstrap reproducibility and posterior class-membership uncertainty of the selected three-class latent class mixed model**

**Panel A. Bootstrap reproducibility of the selected three-class solution.**

| Bootstrap assessment | Result |
|---|---|
| Bootstrap replicates with a usable fitted solution | 500/500 (100%) |
| Stage-2 optimization escalation | 95/500 (19.0%) |
| Stage-3 escalation | 12/500 (2.4%) |
| Optimization resolved under staged criteria | 491/500 (98.2%) |
| Optimization unresolved after the final search | 9/500 (1.8%) |
| Median overall ARI | 0.823 |
| Median Jaccard, Class 1 | 0.932 |
| Median Jaccard, Class 2 | 0.796 |
| Median Jaccard, Class 3 | 1 |
| Median Class-3 recall | 1 |
| Bootstrap replicates retaining all six reference Class 3 participants | 98.20% |

**Panel B. Posterior class-membership uncertainty and bootstrap retention**

| LCMM class | N | Median max posterior probability | ≥0.80 | ≥0.90 | Median normalized entropy | Median bootstrap same-class frequency |
|---|---|---|---|---|---|---|
| Class 1 | 55 | 0.997 | 96.4% | 92.7% | 0.019 | 0.908 |
| Class 2 | 19 | 0.962 | 68.4% | 57.9% | 0.150 | 0.952 |
| Class 3 | 6 | 1.000 | 100.0% | 100.0% | 0.001 | 0.997 |

Panel A summarizes participant-level bootstrap reproducibility. Availability of a usable fitted solution was distinguished from optimization resolution under the staged criteria described in Supplementary Methods, "Bootstrap resampling and optimization." All successful replicates were included in the ARI, Jaccard, recall, and participant-level retention summaries, including those with unresolved optimization. Panel B summarizes membership uncertainty for the selected three-class LCMM. Maximum posterior probability is the highest posterior class-membership probability for each participant. Proportions ≥0.80 and ≥0.90 are the within-class proportions of participants meeting those descriptive probability thresholds. Normalized entropy ranges from 0 to 1, with lower values indicating less posterior class-membership uncertainty. Bootstrap same-class frequency is the proportion of successful bootstrap fits assigning each participant to the reference class, summarized by the median within each reference class. Class 3 comprised six participants. High reproducibility of class assignment should not be interpreted as evidence of precise or broadly generalizable Class 3 clinical or model-parameter estimates. Abbreviations: ARI, adjusted Rand index; LCMM, latent class mixed model.

**Supplementary Table S6. Sensitivity analyses for posterior-propagated associations with the model-derived recovery parameters.**

**Panel A. Sensitivity to the scale used for estimated initial severity (α).**

| Model / Characteristic | Outcome | Natural-scale α specification, median difference (95% CrI) | Model-internal α specification,median difference (95% CrI) |
|---|---|---|---|
| Core: Age, +10 years | r | −0.005 (−0.033 to 0.009) | −0.005 (−0.033 to 0.009) |
| Core: TMT-A, +1 SD worse | r | −0.003 (−0.038 to 0.017) | −0.003 (−0.037 to 0.016) |
| Core: Age, +10 years | τ | 4.1 (−5.5 to 17.5) days | 4.1 (−5.9 to 17.7) days |
| Core: TMT-A, +1 SD worse | τ | 11.9 (−2.7 to 32.9) days | 11.9 (−2.6 to 32.4) days |
| Anatomic: TMT-A, +1 SD worse | τ | 9.7 (−4.0 to 30.4) days | 9.9 (−4.0 to 29.9) days |
| Anatomic: Cerebellum-only vs other | τ | 17.9 (−8.1 to 44.5) days | 17.7 (−8.1 to 45.2) days |
| Functional: TMT-A, +1 SD worse | τ | 7.3 (−5.5 to 24.8) days | 7.3 (−5.8 to 25.1) days |
| Functional: BBS, +10 points | τ | −15.2 (−27.3 to −5.2) days | −15.3 (−27.1 to −5.1) days |

**Panel B. Additional cognition and rehabilitation sensitivity analyses.**

| Sensitivity analysis / Characteristic | Difference in *r*, median (95% CrI) | Difference in τ, days, median (95% CrI) |
|---|---|---|
| Cognition: FAB, +1 SD | 0.001 (−0.023 to 0.024) | −4.5 (−16.3 to 8.7) |
| Rehabilitation: TMT-A, +1 SD worse | −0.003 (−0.038 to 0.018) | 12.7 (−2.5 to 33.6) |
| Physical therapy exposure, per +10 min per PT treatment day | 0.000 (−0.015 to 0.014) | −1.6 (−10.2 to 7.3) |

Values are posterior medians and 95% credible intervals (CrIs). Panel A compares the primary natural-scale α specification with the model-internal α sensitivity specification; α coefficients are not compared because the scales differ. Model-internal α denotes the initial-severity predictor $a_i = \text{logit}(\alpha_i/40)$. Panel B shows the focal results from analyses replacing TMT-A with FAB and additionally adjusting the core model for physical therapy duration; α, age, and sex were included as adjustment covariates but are not repeated for brevity. Natural-scale average marginal differences were calculated at each propagated draw by inverse-transforming the regression linear predictors using the logistic function for $r$ and $\exp(\cdot) + 0.142$ for $\tau$. For continuous covariates, the stated increase was applied to each participant's value; binary covariates were contrasted between 0 and 1, holding the other covariates fixed. The resulting participant-specific contrasts were averaged across all 80 participants. Posterior medians and 95% CrIs were obtained from the median and the 2.5th and 97.5th percentiles of the 4,000 draw-wise averages. These contrasts were based on inverse-transformed linear predictors, without averaging over the secondary-regression residual distribution. VIFs were evaluated across 250 selected primary-model posterior draws for the core, anatomic, and functional models using natural-scale $\alpha$. The maximum VIF was 2.38. Physical therapy exposure was expressed as mean minutes per PT treatment day during hospitalization, using the same definition as in Table 1.

BBS, Berg Balance Scale; FAB, Frontal Assessment Battery; SARA, Scale for the Assessment and Rating of Ataxia; TMT-A, Trail Making Test Part A; VIF, variance inflation factor.

**Supplementary Table S7. MCMC sampling diagnostics for the primary Bayesian nonlinear mixed-effects model.**

| Parameter | R-hat | Bulk ESS | Tail ESS | MCSE of posterior mean | MCSE / posterior SD (%) |
|---|---|---|---|---|---|
| Random-intercept SD: logit($\alpha$/40) | 1.000 | 3,220 | 4,930 | 0.00185 | 1.75 |
| Random-intercept SD: logit(r) | 1.004 | 1,083 | 2,252 | 0.00873 | 2.94 |
| Random-intercept SD: log($\tau - 0.142$) | 1.003 | 1,943 | 3,525 | 0.00249 | 2.28 |
| Population-level intercept: logit($\alpha$/40) | 1.000 | 1,798 | 3,387 | 0.00330 | 2.36 |
| Population-level intercept: logit(r) | 1.001 | 3,074 | 3,749 | 0.00540 | 1.88 |
| Population-level intercept: log($\tau - 0.142$) | 1.001 | 1,833 | 3,050 | 0.00357 | 2.35 |
| Residual SD | 1.000 | 2,350 | 4,609 | 0.00180 | 2.06 |

Sampling diagnostics are reported for the population-level intercepts, participant-level random-effect standard deviations, and residual standard deviation of the primary Bayesian nonlinear mixed-effects model. The model was estimated using four MCMC chains with 4,000 iterations per chain, including 2,000 warm-up iterations, yielding 8,000 post-warm-up draws. Bulk and tail effective sample sizes (ESSs) summarize sampling efficiency in the central and tail regions of the posterior distributions, respectively. MCSE denotes the Monte Carlo standard error of the posterior mean, computed using posterior::mcse_mean() from ordered post-warm-up draws with chain structure retained and reported on the same scale as the corresponding parameter. The final column reports 100 × MCSE/posterior SD. MCSE quantifies finite-simulation error rather than posterior uncertainty or practical identifiability. R-hat values are rounded to three decimal places, ESSs to the nearest integer, MCSEs to five decimal places, and MCSE/posterior-SD percentages to two decimal places.  ESS, effective sample size; MCSE, Monte Carlo standard error; MCMC, Markov chain Monte Carlo; SD, standard deviation.

**Supplementary Table S8. Exploratory region-of-practical-equivalence (ROPE) assessment of posterior-propagated associations with $r$ and $\tau$.**

**Panel A. Core secondary model.**

| Clinical characteristic | Outcome | Posterior median (95% CrI) | Exploratory ROPE | P below ROPE | P within ROPE | P above ROPE |
|---|---|---|---|---|---|---|
| Initial severity α, per +5 SARA points | r | −0.001 (−0.019 to 0.012) | −0.00655 to +0.00655 | 22.3% | 69.0% | 8.6% |
| Initial severity α, per +5 SARA points | τ (days) | 0.8 (−7.5 to 10.8) | −6.18 to +6.18 days | 4.9% | 82.7% | 12.4% |
| Age, per +10 years | r | −0.005 (−0.033 to 0.009) | −0.00655 to +0.00655 | 43.5% | 51.0% | 5.5% |
| Age, per +10 years | τ (days) | 4.1 (−5.5 to 17.5) | −6.18 to +6.18 days | 1.7% | 62.6% | 35.6% |
| Male vs female | r | 0.001 (−0.040 to 0.046) | −0.00655 to +0.00655 | 29.6% | 33.1% | 37.3% |
| Male vs female | τ (days) | 1.6 (−22.1 to 26.7) | −6.18 to +6.18 days | 25.3% | 39.6% | 35.1% |
| TMT-A, per +1 SD in log completion time | r | −0.003 (−0.038 to 0.017) | −0.00655 to +0.00655 | 38.6% | 47.0% | 14.4% |
| TMT-A, per +1 SD in log completion time | τ (days) | 11.9 (−2.7 to 32.9) | −6.18 to +6.18 days | 0.7% | 22.7% | 76.6% |

**Panel B. Functional sensitivity model: additional BBS contrast**

| Clinical characteristic | Outcome | Posterior median (95% CrI) | Exploratory ROPE | P below ROPE | P within ROPE | P above ROPE |
|---|---|---|---|---|---|---|
| BBS, per +10 points | r | 0.003 (−0.015 to 0.027) | −0.00655 to +0.00655 | 10.6% | 53.2% | 36.1% |
| BBS, per +10 points | τ (days) | −15.2 (−27.3 to −5.2) | −6.18 to +6.18 days | 96.1% | 3.9% | 0.0% |

Values are based on 4,000 posterior-propagated draws. The exploratory ROPE was defined as ±0.10 of a reference between-participant standard deviation on the natural parameter scale. The reference standard deviation was the posterior median, across primary-model draws, of the sample standard deviation across the 80 participants; this yielded ROPE bounds of ±0.00655 for r and ±6.18 days for τ. P below ROPE, P within ROPE, and P above ROPE are the proportions of propagated draws below, within, and above these bounds, respectively. The ROPE is used as a relative-magnitude sensitivity analysis and is not a validated clinical-equivalence threshold. The functional sensitivity model included the core covariates plus baseline BBS; Panel B reports only the added BBS contrast to avoid duplicating the core covariates. CrI, credible interval; BBS, Berg Balance Scale; SARA, Scale for the Assessment and Rating of Ataxia; TMT-A, Trail Making Test Part A.
